# High-resolution within-sewer SARS-CoV-2 surveillance facilitates informed intervention

**DOI:** 10.1101/2021.05.24.21257632

**Authors:** Katelyn Reeves, Jennifer Liebig, Antonio Feula, Tassa Saldi, Erika Lasda, William Johnson, Jacob Lilienfeld, Juniper Maggi, Kevin Pulley, Paul J. Wilkerson, Breanna Real, Gordon Zak, Jack Davis, Morgan Fink, Patrick Gonzalez, Cole Hager, Christopher Ozeroff, Kimngan Tat, Michaela Alkire, Claire Butler, Elle Coe, Jessica Darby, Nicholas Freeman, Heidi Heuer, Jeffery R. Jones, Madeline Karr, Sara Key, Kiersten Maxwell, Lauren Nelson, Emily Saldana, Rachel Shea, Lewis Salveson, Kate Tomlinson, Jorge Vargas-Barriga, Bailey Vigil, Gloria Brisson, Roy Parker, Leslie A. Leinwand, Kristen Bjorkman, Cresten Mansfeldt

## Abstract

To assist in the COVID-19 public health guidance on a college campus, daily composite wastewater samples were withdrawn at 20 manhole locations across the University of Colorado Boulder campus. Low-cost autosamplers were fabricated in-house to enable an economical approach to this distributed study. These sample stations operated from August 25^th^ until November 23^rd^ during the fall 2020 semester, with 1,512 samples collected. The concentration of SARS-CoV-2 in each sample was quantified through two comparative reverse transcription quantitative polymerase chain reactions (RT-qPCRs). These methods were distinct in the utilization of technical replicates and normalization to an endogenous control. (1) Higher temporal resolution compensates for supply chain or other constraints that prevent technical or biological replicates. (2) The endogenous control normalized data agreed with the raw concentration data, minimizing the utility of normalization. The raw wastewater concentration values reflected SARS-CoV-2 prevalence on campus as detected by clinical services. Overall, combining the low-cost composite sampler with a method that quantifies the SARS-CoV-2 signal within six hours enabled actionable and time-responsive data delivered to key stakeholders. With daily reporting of the findings, wastewater surveillance assisted in decision making during critical phases of the pandemic on campus, from detecting individual cases within populations ranging from 109 to 2,048 individuals to monitoring the success of on-campus interventions.

**Synopsis:** Tracking SARS-CoV-2 in on-campus wastewater informs and monitors public health decisions and actions.

**TOC/Abstract Art:** 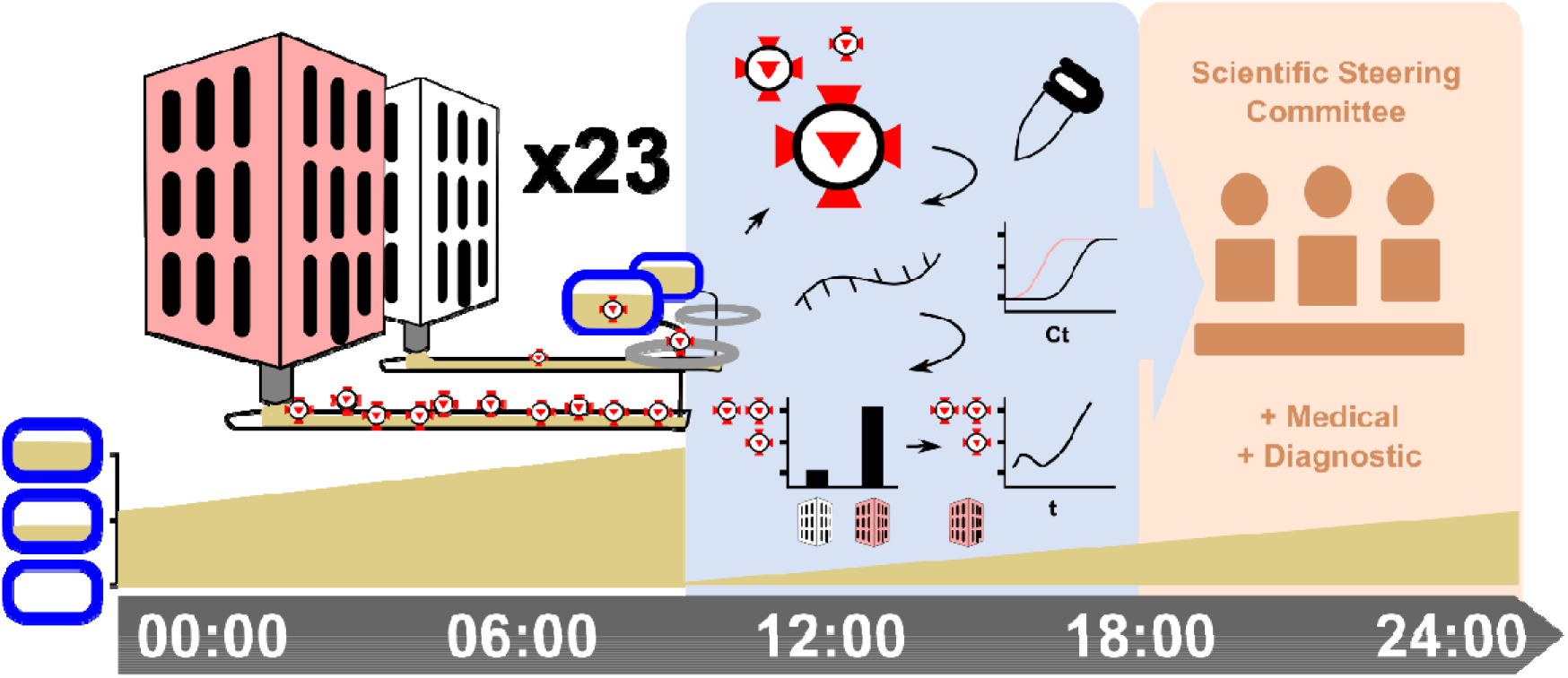

## Introduction

On March 11, 2020, the World Health Organization (WHO) declared coronavirus disease 2019 (COVID-19) a pandemic (1). As of May 20, 2021, 165 million confirmed cases have resulted in over three million deaths (2). Clinical testing of individuals is crucial for identifying infected persons, understanding infection prevalence, and containing the disease, but supply chain limitations and logistical challenges limit clinical testing capacity. Testing is therefore generally reserved for individuals either showing symptoms or likely exposed to the disease (3). However, the etiologic agent responsible for COVID-19, severe acute respiratory syndrome coronavirus 2 (SARS-CoV-2), has a significant asymptomatic percentage (with some estimates of 50%) (4)-(6) and can be transmitted by pre-symptomatic and asymptomatic persons (7)-(11). Further, symptoms may take up to two weeks to develop post-infection (12)-(14), and even when symptomatic, individuals may not self-report. As a result, clinical testing alone fails to identify many infected individuals before they transmit the disease to others and under-represents caseload numbers utilized by officials to inform public health directives. The need to address these shortcomings with a supplementary epidemiological tool was recognized early in the pandemic with a global collaborative of researchers advocating for wastewater-based epidemiology (WBE) (15).

WBE efficiently and non-invasively monitors community metrics by sampling generated wastewater and screening for chemical and biological entities, with previous success demonstrated in tracking community drug use (16),(17) and poliovirus circulation (18)-(20). Wastewater networks can be sampled at points at which discharges from community members have combined, aggregating a semi-anonymous signal representative of the upstream community. Analyzing aggregated wastewater for SARS-CoV-2 RNA therefore provides an opportunity to test entire communities within a single sample. Moreover, as SARS-CoV-2 RNA is present in the feces of both symptomatic (21)-(28) and asymptomatic (29)-(31) COVID-19-infected individuals, wastewater analysis offers insight into infection prevalence unhindered by factors such as symptom onset and the healthcare-seeking behavior of individuals. Further, whereas aggregated testing cannot pinpoint infected individuals, this approach can allow for more effective use of clinical testing resources. For example, WBE can quickly identify the regions and communities with the most infections and allow for the targeted allocation of resources to those “hotspots” for early and comprehensive testing of symptomatic and asymptomatic persons on a localized level (32). Although SARS-CoV-2 RNA fecal shedding behavior has been reported as erratic (33), wastewater represents a complex mixture of all liquid-conveyed waste exiting a premise, including urinary, respiratory, oral, and hygiene-based discharges, compositing multiple potential sources of viral RNA.

An international network of researchers has detected SARS-CoV-2 RNA in their local wastewaters (34)-(43). Their efforts establish proof of concept for WBE in the context of COVID-19 monitoring and work to validate the utility of the approach. These previous studies primarily sampled from wastewater treatment plants (WWTPs)/water resource recovery facilities (WRRFs), efficient locations for obtaining population-level signals. Monitoring the upstream sewer network and sampling at the building- or microsewershed-scale, however, enhances spatial resolution.

The ability to monitor individual buildings (e.g., university residence halls)/small groups of buildings rather than entire municipalities is desirable for more targeted surveillance and response efforts. For example, coupling building-level wastewater sampling two-to-three times per week with clinical testing has been demonstrated as an effective approach at diverse institutions such as the University of Arizona (Arizona, USA) (44), University of North Carolina at Charlotte (North Carolina, USA) (45), and Kenyon College (Ohio, USA) (46). At Hope College (Michigan, USA), Travis et al. (47) implemented a higher collection frequency by sampling from nine on-campus residential zones every weekday. In these campaigns, wastewater samples indicating infection prevalence led to clinical testing of all individuals associated with the flagged buildings/populations (subject to university-specific decision frameworks). All found WBE to be a valuable tool for disease containment, often noting wastewater surveillance’s utility for identifying and isolating asymptomatic individuals. In light of these successes, more experience and guidance are desired to inform further implementation of building-level WBE campaigns (48).

Here, we report the WBE campaign conducted at the University of Colorado Boulder (Colorado, USA). We sampled up to 20 manhole locations seven days per week between August 25th and November 23rd to monitor on-campus residential buildings for the presence of COVID-19. To obtain economical 24-hour composite samples, we designed, assembled, and deployed low-cost autosamplers. We tracked the concentrations of SARS-CoV-2 RNA and control species in the wastewater using reverse transcription quantitative polymerase chain reaction (RT-qPCR) assays following best practices (49) and reported to campus decision makers daily. This campaign was coupled with weekly saliva-monitoring RT-qPCR testing of all asymptomatic on-campus residents (50),(51). The comprehensiveness of both the WBE campaign and the clinical and monitoring testing services provides a unique opportunity to evaluate the effectiveness of building-level wastewater surveillance and its required temporal frequency.

## Materials and Methods

### Sample Locations

Twenty sample locations targeting the wastewater outfall captured within surface-accessible manholes were prioritized to discriminate the SARS-CoV-2 signals originating from the on-campus residential buildings at the University of Colorado Boulder (**Figure 1 a, Supplemental Table 1**). Twenty-three pumps (autosamplers) operated at these locations, with three sites discriminating two flows within a single manhole. Overall, then, 23 flows originating from 20 manhole locations were monitored. Each flow roughly corresponded to an individual residential structure. The university housed over 6,200 students in on-campus residential buildings during the semester, and each site on average accounted for the wastewater generated by 450 residents (range extending from 109 to 2,048 residents), with select residents being monitored at multiple sites. The targeted manholes ranged from approximately 1 to 7 m in depth. To protect the privacy of residents, the presented data was anonymized with a unique label assigned to each residential structure indicating its position and any other residential structure contributing to its associated flow in parentheses: A, B(A), C, D, E(CBA), F, G(FEDCBA), H, I(H), J, K, Admin, L(Admin), M, N, O, P, Q, R, S, and Isolation.

**Figure 1.**
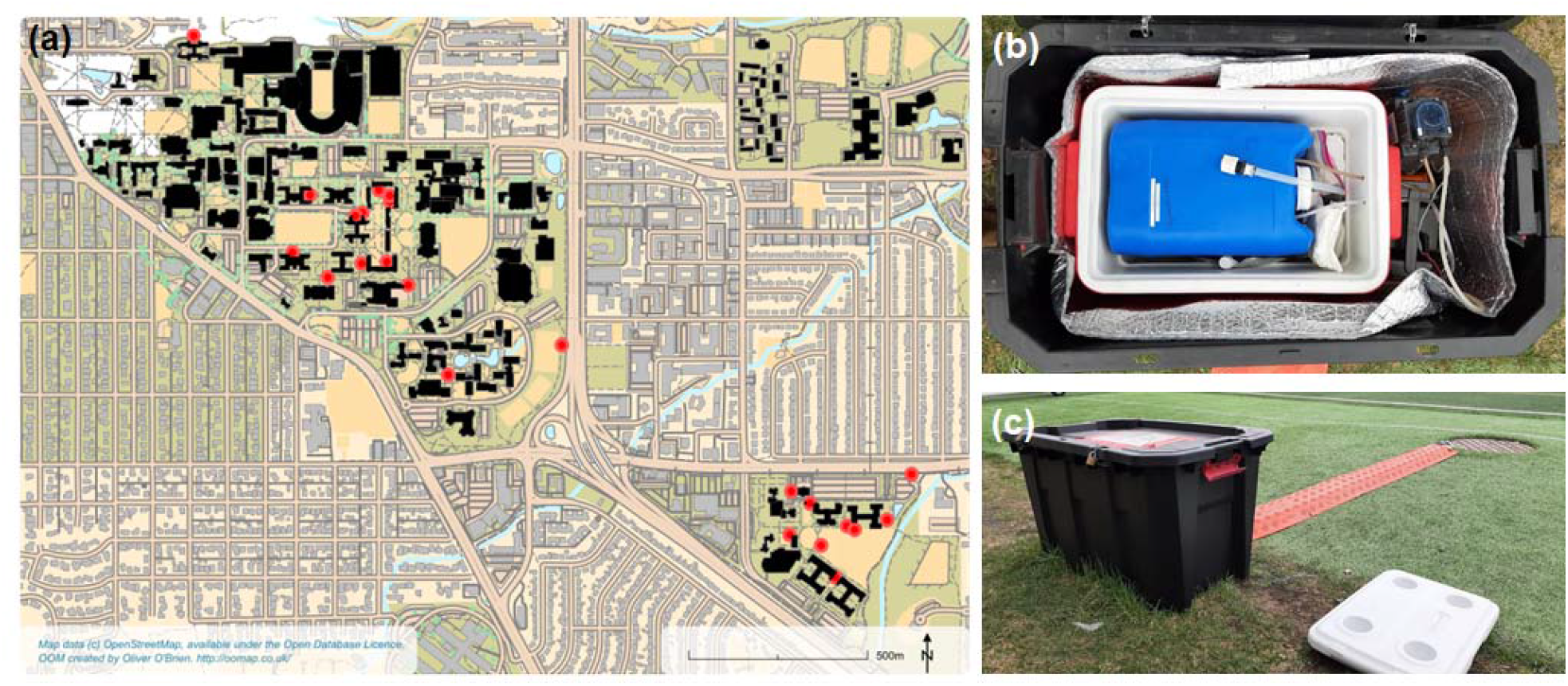
(a) Map of sample locations distributed across the University of Colorado Boulder’s campus. (b) Picture of the internal components of the composite autosampler design. (c) Picture of the composite autosampler in operation.

### Composite Autosampler

The composite autosampler was assembled from readily available materials. The main components were a 24-V Stenner pump (E10VXG; Stenner Pump Company, Jacksonville, FL, USA; designed by DEWCO Pumps, Denver, CO, USA), a 300-Wh portable DC/AC power bank (R300; GoLabs Inc., Carrolton, TX, USA), a 5-gal jerrycan (Uline, Pleasant Prairie, WI, USA), a 9-gal cooler, gel ice packs, insulation, ¼-in. O.D. PVC tubing, and exterior casing (**Figure 1 b, Supplemental Figure 1, Supplemental Table 2**). The samplers were positioned above ground and next to each manhole (**Figure 1 c**). The wastewater inflow and overflow tubing lines were fed through the D-pick of the manhole cover, and the inlet strainer that resided in the underground wastewater stream was constructed from either ¼-in. O.D. copper or ¼-in. O.D. steel tubing, with 0.157-in. holes drilled into the side.

The pumps were continuously operated at approximately 33% of full capacity, each scheduled to draw about 10 L of wastewater per day. The actual wastewater withdrawn was monitored at the time of sample collection by weighing the jerrycan mass with a luggage scale (**Supplemental Figure 2, Supplemental Table 3**). Collection occurred daily between 7 AM and 12 PM, with power banks replaced every 48 hours and ice packs replaced daily when ambient average temperatures exceeded 4°C. Samplers were only turned off for the following three events: a September blizzard, a supply chain disruption in October, and a cold event in October.

### Sample Collection

Two 50-mL subsamples and one 40-mL subsample were collected from each sampler daily. One 50-mL subsample was collected for RNA extraction and viral detection, and the second 50-mL subsample was collected for determination of basic water quality parameters. The 40-mL subsample was collected as a backup sample. Subsamples were poured from the 5-gal jerrycan after swirling the contents. All samples were collected in pre-weighed sterile polypropylene tubes loaded with 500 μL of 10% Tween™ 20 detergent (Thermo Fisher), which served to inactivate infectious agents for worker safety. Samples were stored on ice immediately after collection and during transport back to the laboratory. After sample collection at each sampler, ½-in. O.D. PVC tubing was connected to the jerrycan, fed through the D-pick of the manhole cover, and used to drain excess withdrawn wastewater. Emptied jerrycans were then briefly rinsed with dilute bleach solution and water before being repositioned to collect wastewater over the next 24 hours.

### Sample Processing

All samples were processed in the laboratory the same day as collection. Samples that could not be processed upon immediate arrival at the laboratory were temporarily stored at 4□. Backup and RNA extraction/viral detection samples were processed within 4 hours of arrival; water quality samples were processed within 9 hours of arrival. All samples were spiked with a known amount of bovine coronavirus (Bovilis^®^ Coronavirus; Merck Animal Health, NJ, USA) serving as the internal process control.

#### Water Quality Parameter Samples

The pH (measured with accumet™ AB150 pH meter, Thermo Fisher) and total suspended solids (TSS) values of each sample were measured following standard methods (**Supplemental Figures 3, 4**; **Supplemental Tables 4, 5**).

#### RNA Extraction/Viral Detection Samples

Samples collected for the detection of SARS-CoV-2 RNA were weighed and then centrifuged at 4,500 × g for 20 minutes at 4□. Viruses were concentrated from approximately 35 mL of each sample’s supernatant using ultrafiltration pipettes (CP-Select™ using Ultrafiltration PS Hollow Fiber Concentrating Pipette Tips; InnovaPrep, Drexel, MO, USA) following the manufacturer’s guidelines. Concentrate eluted from the ultrafiltration pipettes was captured and weighed in pre-weighed 15-mL tubes. RNA was then extracted from the concentrate using RNA PureLink Mini Kits (Thermo Fisher) according to the manufacturer’s protocol (with the exception that Wash Buffer □ was not used). A 1-μL aliquot was taken from the extracted RNA of each sample and analyzed on a Qubit™ 4 Fluorometer (Q33238, Thermo Fisher) using the High Sensitivity RNA Kit to quantify the total RNA extracted and roughly assess the success of the extraction process (**Supplemental Table 6**).

#### RT-qPCR

Two separate RT-qPCR pipelines were then used to detect and quantify SARS-CoV-2 RNA. The first RT-qPCR pipeline, entitled SURV1, was executed simultaneously with the sampling campaign. Immediately after the RNA extraction step, a 5-μL aliquot of extracted RNA from each sample was combined with 3-μL of RNase-free water and transported on ice to the University of Colorado Boulder’s COVID Surveillance Laboratory. The remaining volume of extracted RNA was stored at -80□. The COVID Surveillance Laboratory tested saliva samples submitted by on-campus residents and employees for the presence of SARS-CoV-2, and their team performed their RT-qPCR multiplex assay on the extracted RNA wastewater samples in addition to processed saliva samples. SURV1 employed the Centers for Disease Control and Prevention (CDC) multiplex assay targeting the SARS-CoV-2 nucleocapsid (N) and envelope (E) genomic regions as well as the human RNaseP transcript **(Supplemental Figures 5, 6**) (52)-(52). From August 28th to September 29th, the N1 primer and probe set was used to detect the nucleocapsid region. After September 30th, the N2 primer and probe set was used instead because of supply availability. Multiple technical replicates were not run. The wastewater samples were analyzed by SURV1 the same day as sample collection.

The second RT-qPCR pipeline, entitled SENB+, was executed in December 2020 after the fall sampling campaign had ended. In this second pipeline, the extracted RNA samples (frozen at -80□) were reevaluated for SARS-CoV-2 RNA using a wastewater-specific RT-qPCR multiplex assay detecting the following targets: SARS-CoV-2 N (N2), SARS-CoV-2 E, the spiked internal control bovine coronavirus, and genogroup II F+ RNA bacteriophage (**Supplemental Table 7**). Genogroup II F+ RNA bacteriophage was targeted to serve as a human fecal indicator (53).

SENB+ RT-qPCR amplifications were performed in 20-μL reactions including 5-μL TaqPath™ One-Step Multiplex Master Mix (Thermo Fisher), 0.015-μL bovine coronavirus and 0.015-μL F+ bacteriophage primer from 200-μM stock solutions, 0.045 μL of each SARS-CoV-2 primer from 200-μM stock solutions, 0.02 μL of each probe from 100-μM stock solutions, 9.68-μL RNase-free water, and 5-μL template RNA. These volumes created 150-nM bovine coronavirus primer, 150-nM F+ bacteriophage primer, and 450 nM of both SARS-CoV-2 primers with 100 nM of each probe in each reaction. Primer concentrations were chosen to limit amplification of bovine coronavirus and F+ bacteriophage RNA. Each run was performed on a QuantStudio 3 (Thermo Fisher) according to the following program: UNG incubation at 25°C for 2 minutes, reverse transcription at 53°C for 10 minutes, polymerase activation at 95°C for 2 minutes, and amplification in 40 cycles of 95°C for 3 seconds (denaturing step) and 60°C for 30 seconds (annealing and elongation step). Reactions were performed in triplicate. Each run included between one and three triplicate negative control reactions (with 5 μL of RNase-free water instead of template RNA) and a ten-fold serial dilution of single-stranded DNA (F+ bacteriophage (5’-TCTATGTATGGATCGCACTCGCGATTGTGCTGTCCGATTTCACGTCTATCTTCAGTCATTGGATTTGGGGTCTTCTGATCCTCTATCTCCAGACTTTGATGGACTTGCCTAC-3’); IDT Technologies, Coralville, IA, USA) and RNA (SARS-CoV-2 (SARS-CoV-2 Jul-28-20 #2; Twist Biosciences, San Francisco, CA, USA) and bovine coronavirus (direct extraction of the Bovilis^®^ Coronavirus quantified using a Qubit 4)) standards for standard curve quantification. The ten-fold dilutions ranged from 10^5^, 10^6^, and 10^6^ copies to 1, 10, and 10 copies of SARS-CoV-2, bovine coronavirus, and F+ bacteriophage standard per reaction, respectively (**Supplemental Figure 7**). The lower standard amount established the limit of quantification (LOQ). The limit of detection (LOD) was set at amplification occurring before the 40th cycle and above the background amplifications in the extraction blank (**Supplemental Figure 8**) and negative control reactions. Select runs included an additional standard dilution containing 1.53×10^7^ copies of bovine coronavirus standard, assisting quantification when the bovine coronavirus spike-in amount was increased from approximately 50,000 to 500,000 copies per reaction (**Supplemental Figure 9**).

### SENB+ RT-qPCR Data Quality Control

Any amplification with a Ct value roughly 2 or more cycle numbers different than the Ct value of either of the other two amplifications in its triplicate was excluded from the dataset. Amplifications indistinguishable from background drift were excluded from the dataset. Standard dilutions were excluded from standard curve creation if any amplifications of a target observed in the negative controls on the same plate had a lower Ct value than any one of the target’s three standard dilution amplifications in triplicate. Standard dilutions were also excluded from standard curve creation if their amplifications were greatly displaced from their expected position. More specifically, ten-fold standard dilutions amplified with 100% efficiency should be spaced 3.32 cycle numbers apart. Let n represent the intervals between dilutions such that 10^6^ and 10^5^ dilutions are n = 1 interval apart and 10^6^ and 10^4^ dilutions are n = 2 intervals apart. A standard dilution was excluded from standard curve creation if the average Ct of its amplifications was either less than 2n or greater than 5n cycle numbers apart from the average Ct of the amplifications of the closest higher dilution accepted and positioned n before it.

### Data Normalization

SARS-CoV-2 data from SURV1 was normalized by subtracting the RNaseP Ct value from the SARS-CoV-2 E Ct value because these values are logarithmic in nature. The N gene was utilized to confirm trends. Data from SENB+ was processed by calculating copies per liter of wastewater using the recorded masses of sample concentrated and eluted (**Supplemental Table 10**) and the following equation:

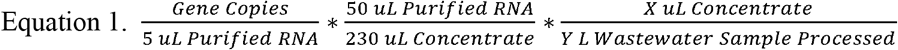

The bovine coronavirus and F+ bacteriophage signals were used to track sample variability but not to transform the concentration of SARS-CoV-2 RNA. The bovine coronavirus recovery efficiency was determined by comparing the RT-qPCR-obtained concentration value to the extracted RNA concentration of the spiked-in control. Throughout the campaign, the recovery efficiency averaged 53 +/- 30% S.D. October 2nd samples are masked from this analysis because they were frozen prior to extraction and a key intermediate weight was not recorded.

### Incorporation of Medical Services and Isolation Space Utilization Data

On-campus medical services in Wardenburg Health performed nasal-swab Lyra® Direct SARS-CoV-2 assays (Quidel Corporation, San Diego, CA, USA) to confirm suspected cases within the community. These data are considered as “positive detections” within the residential structures, and the date of each positive is used to denote the case (though that date is not the date of actual infection) (**Supplemental Table 11**). Isolation space utilization tracks the number of beds in designated isolation spaces occupied on a given day (**Supplemental Table 12**).

## Results and Discussion

### Performance of the Composite Samplers

In general, the composite samplers performed well, reliably withdrawing sample mass. The design achieved the objectives and provided an economical sampling unit. Additionally, if a source of electricity is near the sample point, then the cost decreases with removing the necessity of the power bank. Throughout the campaign, concerns were noted over (1) leakage through the small sampling port on the jerrycan and (2) the inlet strainer either clogging or being knocked offline because of toilet paper accumulation during low-flow conditions. To prevent further leakage, a short PVC tube was epoxied to the small sampling port and positioned such that the free end of the tube sat (with a removable cap) above the jerrycan. Several redesigns of the inlet strainer suffered similar issues as the primary design, exacerbated by the increasing prevalence of “flushable” wipes. Manually unclogging and redeploying the inlet strainers remained the primary maintenance demand. Future surveillance campaigns may consider more permanent modifications to the flow path to enable ease of sample collection.

### Dataset Summary

Prior to resumption of on-campus activities, incoming on-campus residents were required to test five days prior to the scheduled move-in (August 17^th^ -21^st^), establishing the baseline. An initial surge in SARS-CoV-2 RNA wastewater concentrations was detected at the beginning of the campaign (**Figure 2**). This event fell two weeks after the Labor Day holiday in the USA, with many traced large off-campus gatherings. The wastewater concentrations plateaued the week of September 15^th^ and were in decline prior to Boulder County enacting aggressive social distancing policies on September 24^th^ (**Figure 2 a**). That concentrations were already decreasing before these policies were enacted likely reflects the success of on-campus testing, tracing, and isolation efforts. The September 24^th^ orders were enforced until October 13^th^ and prohibited (1) anyone aged 18 to 22 years old in the City of Boulder from engaging in gatherings and (2) residents in 36 nearby off-campus buildings from leaving their place of residence to the maximum extent possible (“stay-at-home” order) (54). Those 36 buildings were identified as likely large-gathering areas. The higher SARS-CoV-2 wastewater concentrations noted on September 27^th^ appear to be a true signal, with both the bovine coronavirus and F+ bacteriophage targets displaying similar abundance ranges to surrounding dates, highlighting the need and utility of multiplexed controls. This peak occurred on a Sunday 24 hours after the identification and isolation of numerous cases at the end of the September surge (**Figure 2 f, h**) and is notable when isolation building inputs are excluded from the wastewater concentration data.

**Figure 2.**
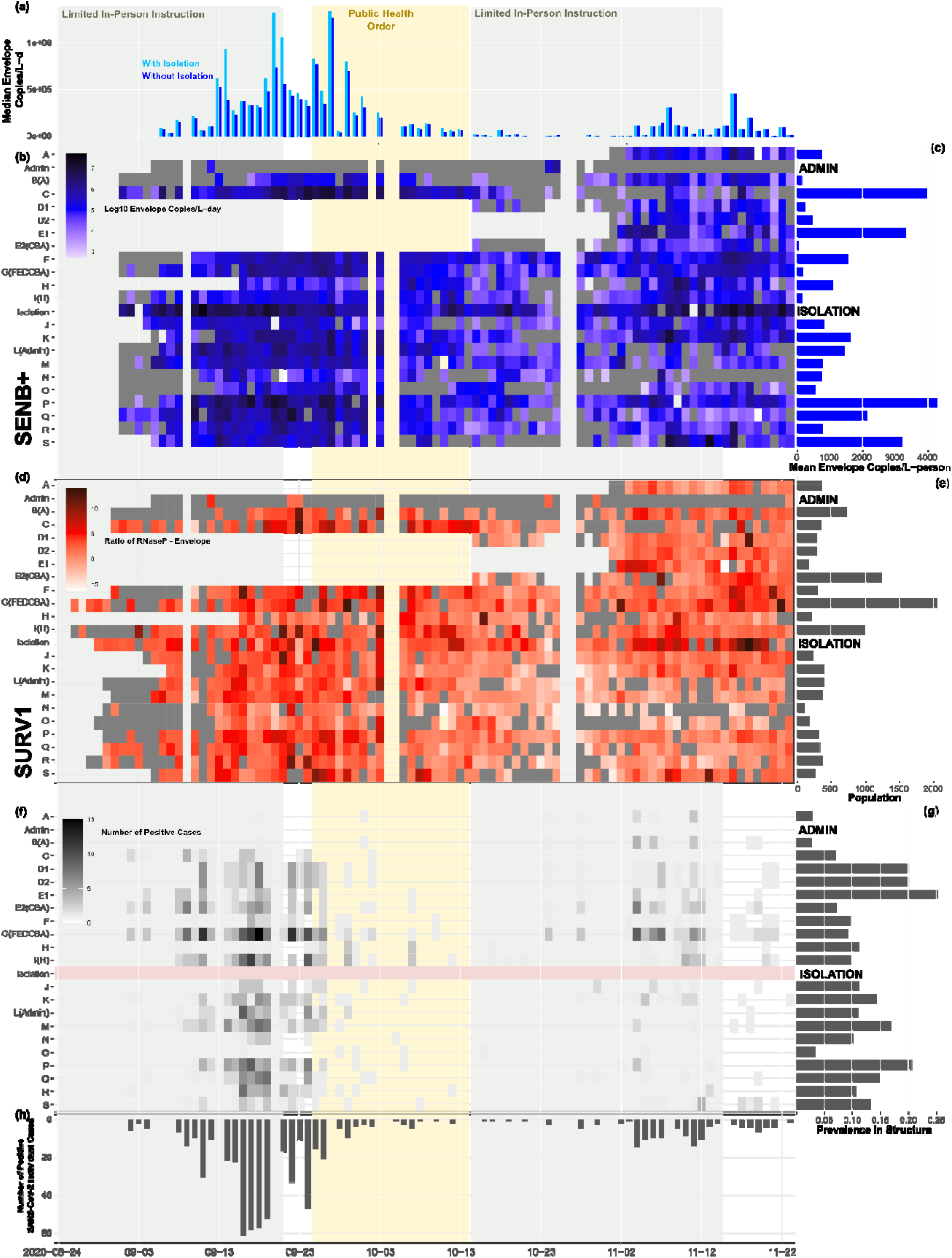
(a) Median SARS-CoV-2 E copies/L wastewater as determined by the SENB+ pipeline. (b) Heatmap displaying the SENB+ SARS-CoV-2 E copies/L on a log scale; grey indicates no detectable amplifications. (c) Per capita average SENB+ SARS-CoV-2 E copies/L over the sampling campaign distributed per sampled wastewater flow, indicating the overall temporal prevalence of SARS-CoV-2 within a single structure. (d) Heatmap displaying the ratio of SARS-CoV-2 E copies/L to the human RNaseP copies/L as determined by the SURV1 pipeline on a log scale; grey indicates no detectable amplifications. SARS-CoV-2 N concentrations confirm the displayed trends (**Supplemental Figure 10**). (e) Population served by each sampler. (f) Heatmap displaying the confirmed medical services positives mapped to each sampler. (g) Prevalence, measured by the total number of SARS-CoV-2 infections detected among a population served by a sampler divided by the total number of that population. (h) Sum of confirmed positives per day.

Well after the expiration of those public health orders, another increase in wastewater concentrations was detected after October 31st (the Halloween holiday in the USA). Clinical services detected fewer cases on-campus during this event as compared to the September event, and this lower prevalence was reflected in the lower SARS-CoV-2 wastewater concentrations. The similar dynamics emphasize a quantitative relationship, not simply a presence or absence of viral RNA correlation, between the SARS-CoV-2 prevalence and the wastewater concentrations. Finally, students vacated campus prior to November 23^rd^ for the scheduled end of in-person instruction. The SARS-CoV-2 wastewater concentration averages taken over the monitoring campaign (**Figure 2 c**) largely reflected on-campus prevalence (**Figure 2 g**) (number of reported infections within a residential structure divided by the initial census data, **Figure 2 e**) when masking those sample sites that were activated later in the semester.

Both the SURV1 data and the SENB+ data reflected the medical services data throughout the campaign (**Figure 2 b, d; Supplemental Tables 8, 9**). Overall, the data from the SURV1 and SENB+ pipelines are consistent, suggesting that a single technical replicate (the SURV1 dataset) is admissible when performing a daily monitoring campaign and when resources become limited resulting from either supply chain disruptions or rapid campaign expansions designed to meet the pace of emerging pandemics. Technical replicates are still recommended when available (the SENB+ dataset), though, to avoid false reporting. Additionally, the SARS-CoV-2 E and N2 targets displayed a linear correlation of 0.97 in the SENB+ data (**Supplemental Figure 10**), confirming that both are suitable to track the prevalence of the virus. However, especially considering the detection of SARS-CoV-2 variants in wastewater (55), continuing to track multiple locations along the SARS-CoV-2 genome provides critical robustness against false negative and positive events. The quantitative range of the predicted concentrations (in terms of genome copies per liter of wastewater) was also similar for both targets. The E target, however, reported fewer non-detects and thus displayed a higher sensitivity than the N2 target. The E target was therefore utilized as the primary dataset considered daily, with the N2 target serving a confirmatory function.

Relating the concentration of SARS-CoV-2 RNA in wastewater to the medical services data was importantly influenced by the isolation strategy used on campus. The majority of positive individuals received temporary housing in the primary isolation building up until September 18^th^ and after October 6^th^ and were assigned to a secondary building between September 18^th^ and October 5^th^ (**Figure 3**). However, select students were allowed to isolate in place (structures A and B). Isolation in these alternate structures complicated the signal in their associated wastewater flows as well as in the combined G(FEDCBA) flow (the E2(CBA) flow is not noted given the sampler serving that structure primarily operated after October 5^th^). Additionally, such combined flows bias the median data (**Figure 2 a**), with the contribution of a single infection potentially detected in, at the maximum, four sites. This complication emphasizes the importance of quantifying the signal for these locations rather than relying on binary presence/absence of virus determinations. Quantification enables the detection of temporal trends such as increasing SARS-CoV-2 RNA concentrations above the expected baseline. Additionally, students do not proceed through the entire course of infection profiled within a given residence and shift their contribution to the isolation structures (**Figure 3**). A similar behavior must be accounted for within broader wastewater networks, in which movement of individuals seeking medical services and requiring longer stays within hospitals and long-term care facilities potentially decouples the wastewater signal from the served residential units.

**Figure 3.**
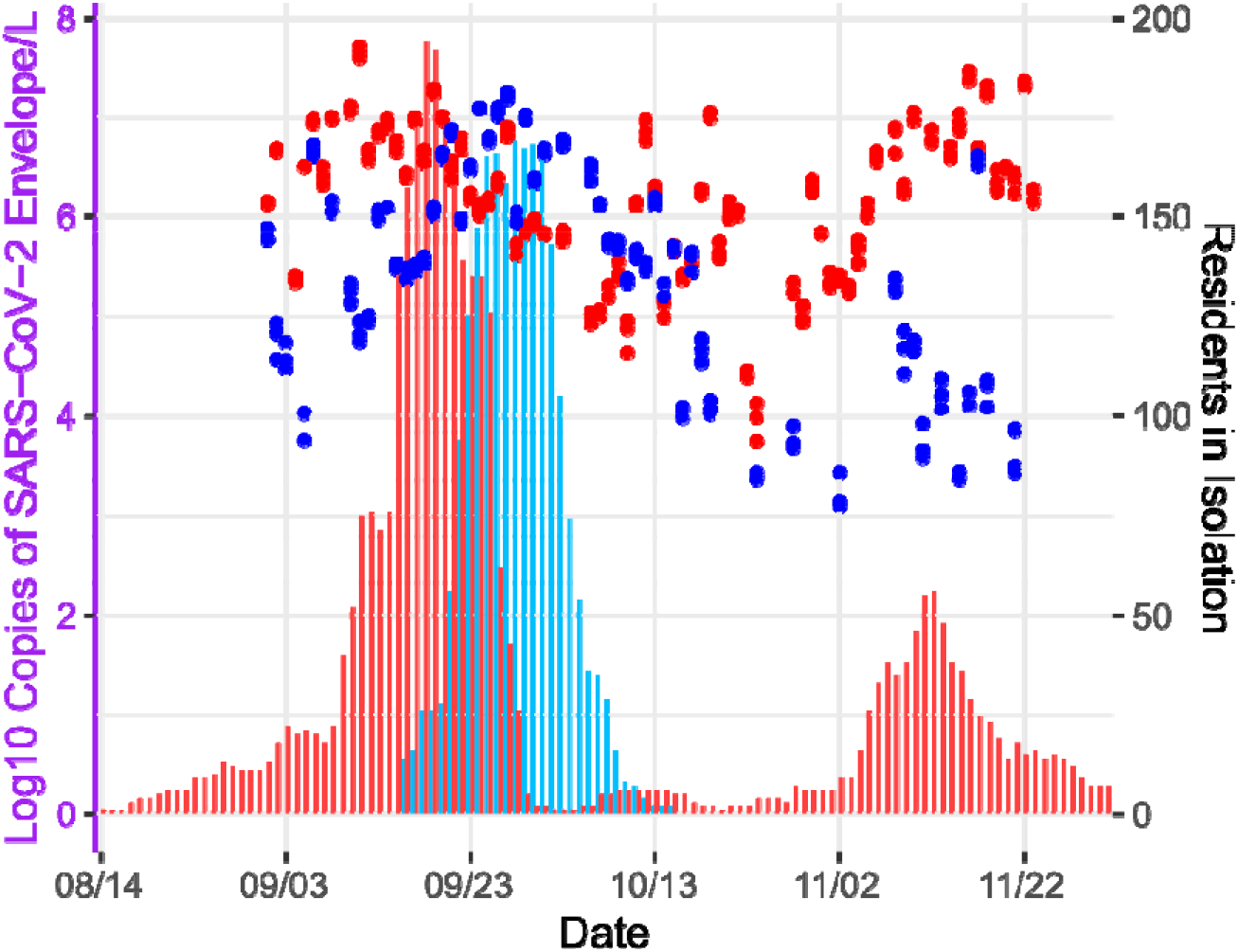
Residency reported (bars) versus SENB+ SARS-CoV-2 E copies/L wastewater (points) detected for the primary (red) and backup (C[Backup Isolation], blue) isolation structures.

The primary isolation building maintained a unique wastewater in which the flow did not represent a fluctuating proportion of infected individuals amongst non-infected individuals. The building was instead occupied entirely by individuals progressing through the course of the viral infection, remaining empty otherwise. The wastewater concentrations from this building peaked in mid-September and again in mid-November at approximately 10^7^ SARS-CoV-2 copies/L wastewater (**Figure 3**). These peaks resulted from the co-occurrence of disease progression and virus/viral RNA shedding in stool, explaining the two peaks’ nearly identical wastewater concentrations despite substantially different infected resident numbers. In this building, the viral wastewater inputs of a smaller number of infected individuals were not more diluted by the inputs of a corresponding larger proportion of non-infected residents; the isolated individuals’ wastewater mixed only with idle plumbing and appliance flows. The peaks noted over October reflect the progression of viral shedding from individual contributions. Although this value will vary with the underlying characteristics of the idle flow emanating from each building, from the presented data, the expected maximum concentrations of detectable SARS-CoV-2 within domestic wastewater in the USA should be near 10^7^ genome copies/L. Considering that individuals in residences are expected to produce between 100-250 L of wastewater per day, the maximum shedding per person is on the order of 10^10^ SARS-CoV-2 genome copies/day, in agreement with Schmitz et al. (56). This number additionally aligns with the upper-end of identified fecal concentration ranges, suggesting that individuals within these structures likely produce between 100-1,000 mL of feces per day (5×10^3^–10^7.6^ copies/mL feces) (57). The campus additionally relied on a secondary isolation building during the peak of infections in September (**Figure 3**). Notably, this structure displayed a similar maximum SARS-CoV-2 wastewater concentration.

The concentration of fecal matter becomes more critical when considering wider communities with industrial, infiltration, and other diluting contributions to wastewater. In initial attempts to normalize to the varying concentrations of fecal matter within the wastewater samples, the genogroup II F+ bacteriophage was selected as an internal reference marker for the fall campaign to align with other sampling efforts ongoing within Colorado. At the micro-sewershed level, the F+ bacteriophage signal displayed inconsistent geographical and temporal trends (**Figure 4**). Select sites (e.g., R, Q, and O) displayed consistently low signals, within the range of 10^4^ to 10^6^ copies/L, whereas other sites (e.g., G(FEDCBA), J, and L(Admin)) displayed signals often approaching 10^9^ copies/L. Even more concerning, sites such as C, F, H, M, and N display inconsistent temporal trends, fluctuating over five orders of magnitude during the fall campaign. These shifts potentially result from changes in resident diet or interpersonal fluctuations in the gut virome (58). The F+ bacteriophage was therefore replaced by the pepper mild mottle virus (PMMoV) for the spring 2021 monitoring campaign (59).

**Figure 4.**
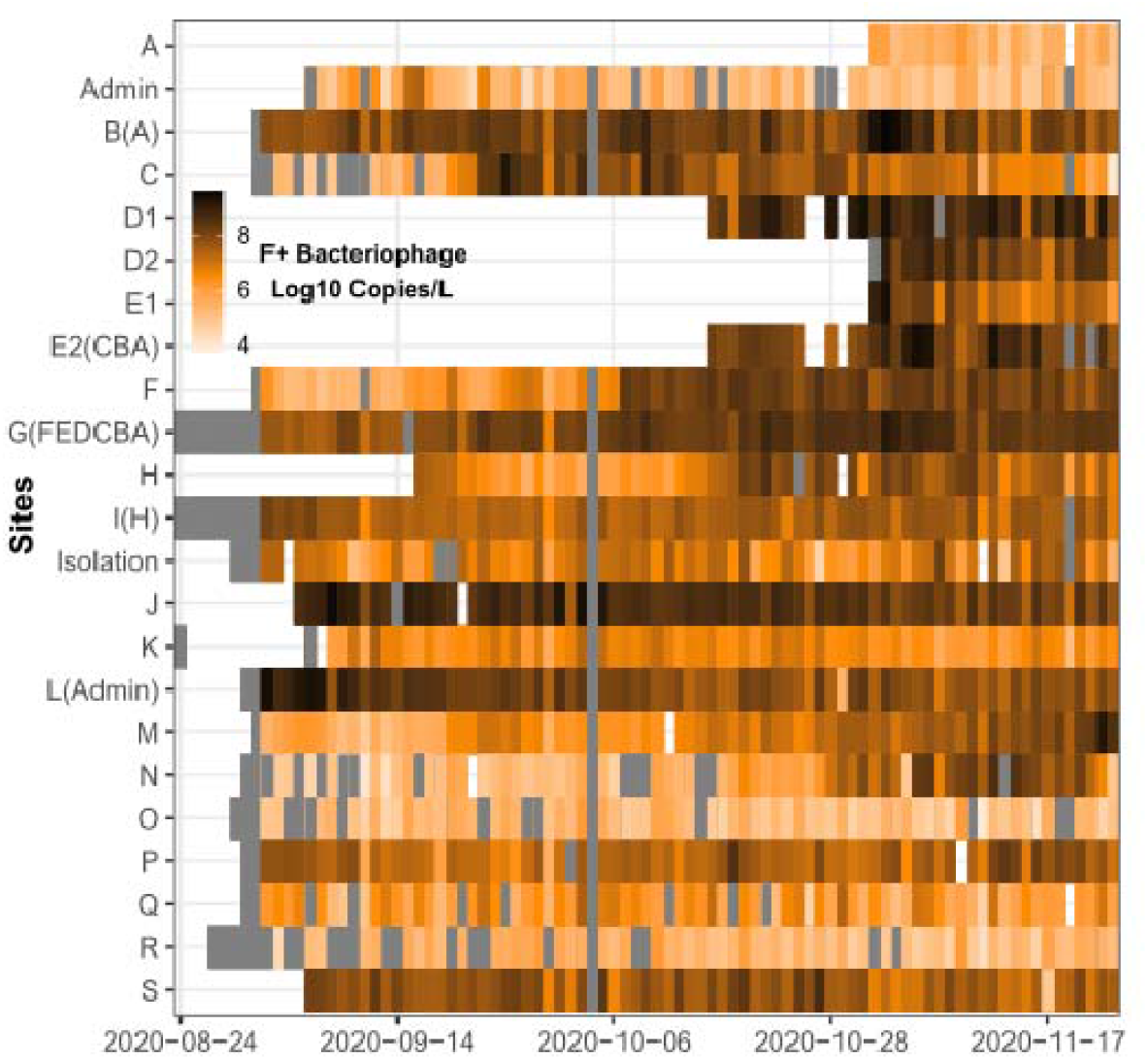
Heatmap of the F+ bacteriophage copies/L wastewater detected on a log scale. Darker shades of orange indicate higher concentrations, with grey indicating no detectable amplifications.

### Utility and Consideration of the Data

Throughout the fall monitoring campaign, the interpretation and utility of the data varied with the prevalence of SARS-CoV-2 within the community. Considering six scenarios in which the concentration of SARS-CoV-2 is (1) absent, (2) low and stable, (3) low and increasing, (4) high and increasing/stable, (5) high and decreasing, and (6) decreasing to absent, the daily monitoring campaign provided varying levels of support to the pandemic response. The utility as an early warning signal is primarily experienced in scenarios (1), (2), and (3), in which early detections are the most critical for preventing or halting community spread. This prevention requires a robust and well-connected testing, contact tracing, and isolation infrastructure. When entering either scenario (4) or (5), the primary utility in WBE is in monitoring the effectiveness of public health intervention strategies employed. The fall campaign provided an example, in which the peak in SARS-CoV-2 wastewater concentrations occurred before the social distancing order imposed by the county. Therefore, the on-campus mechanism of testing, tracing, and isolation was demonstrated as effective prior to a more robust and stringent social distancing order being put in-place. This monitoring better equips public health officials to determine appropriate responses with the infrastructure at hand, with more stringent control measures likely leading to migration from campus and potentially transporting viral infections further abroad and/or allowing reentry of the virus from broader community-acquired infections when social distancing requirements ease. After the public health orders were enacted in Boulder, Wi-Fi connections within residence halls decreased by 33%. Additionally, clinical testing data later in the semester identified cases with low viral loads without an active infection, highlighting cases in which progression through the disease profile occurred off campus. Finally, during scenario (6), wastewater data also effectively monitors individuals as they exit the infectious period but may still be shedding viral RNA. On campus, students were permitted to leave the isolation structure and return to their residences after ten days. These reentry events could be detected in the wastewater (e.g., see site O, **Figure 2**). Reentries thus must also be taken into consideration to prevent shifts in policy based on a true detected signal that is not reflective of a case of concern.

## Conclusions

With the wide range of fluctuations in daily habits surrounding toilet flushes and personal hygiene behaviors combined with rapid changes in viral loads, daily monitoring becomes critical to track the prevalence of pathogens within building-scale wastewater. The presented monitoring campaign is distinguished by high temporal and geographical resolution over a university campus. However, a tradeoff emerges considering the commitment of resources versus the action items taken surrounding the usage and monitoring of the data. The demonstrated monitoring campaign informed on the emergence of likely new infections within given residential structures, notably during the first two weeks of operation, and the effectiveness of on-campus interventions. The utility of these data relied on being in concert with robust medical services and monitoring testing data, providing the ability to translate from community monitoring to intervention. Across university campuses scattered globally, and reported within this study, the utility of wastewater monitoring to support public health has been demonstrated. This study concluded that (1) economical solutions are readily assembled for operating composite samplers, (2) daily samples enable informed decisions and monitoring of the success of interventions on-campus, and (3) wastewater data provides substantial and unique benefit when surveying community health at multiple stages in a disease outbreak. Combined, wastewater monitoring provides a flexible and effective public-health technique when deployed at the building-level scale.

## Supporting information

Supplemental Script

## Data Availability

All data associated with this manuscript is attached within the supplemental materials.

## Acknowledgements

We thank Brian Graham, Gary Low, Jonathan Akins, Chris Busch, and David Lawson at the University of Colorado Boulder in assisting with site selection and deployment. Holly Gates-Mayer (University of Colorado Boulder) assisted with the safety assessment and PPE assignment. We thank Susan De Long (Colorado State University), Carol Wilusz (Colorado State University), and Rebecca Ferrell (Metropolitan State University of Denver) for assistance in the development of the in-house extraction and molecular methods. Additionally, John Spear (Colorado School of Mines), Tzahi Cath (Colorado School of Mines), Kari Sholtes (Colorado Mesa University), and Keith Miller (University of Denver) provided valuable feedback into the design and operation of the campaign. Funding for this project was provided by the CARES Act, administered by the Office of Equity, Compliance, and Integrity at the University of Colorado Boulder, and overseen by a collaborative effort through the Scientific Steering Committee, notably Matt McQueen, Jennifer McDuffie, Mark Kavanaugh, and Melanie Parra. We also wish to thank the broader wastewater surveillance community for rapid collaborations and sharing of ideas and techniques and the operators, engineers, and managers at the Boulder Water Resource Recovery Facility. Finally, we thank all of the students who resided on campus.

## Supporting Information

The supporting information is available free of charge at: [HTML]

The supporting tables contain the sample location details, component list for the sampler design, daily mass of wastewater collected, daily wastewater pH, daily wastewater total suspended solids (TSS), concentration of total RNA extracted, primers and probes used in the SENB+ multiplex, SURV1 RT-qPCR Ct data, SENB+ RT-qPCR concentration values, processing data required to back-calculate to copies per L of wastewater, medical services determined positives per manhole, and residency within isolation structures. The supporting figures contain the schematic of the sampler design, daily wastewater mass, daily wastewater pH, daily wastewater TSS, processing controls, standard curves, extraction blanks, bovine coronavirus recovery, and comparison between the nucleocapsid (N) and envelope (E) targets. A supplemental compressed file contains the required script and files to generate the presented data analysis and statistical graphics.

## References

(1) WHO Director-General’s opening remarks at the media briefing on COVID-19 - 11 March 2020 https://www.who.int/director-general/speeches/detail/who-director-general-s-opening-remarks-at-the-media-briefing-on-covid-19---11-march-2020 (accessed Dec 10, 2020).

(2) World Health Organization (WHO). WHO Coronavirus (COVID-19) Dashboard https://covid19.who.int (accessed May 21, 2021).

(3) Centers for Disease Control and Prevention (CDC). COVID-19 and Your Health https://www.cdc.gov/coronavirus/2019-ncov/testing/diagnostic-testing.html (accessed May 7, 2021).

(4) Mizumoto, K.; Kagaya, K.; Zarebski, A.; Chowell, G. Estimating the Asymptomatic Proportion of Coronavirus Disease 2019 (COVID-19) Cases on Board the Diamond Princess Cruise Ship, Yokohama, Japan, 2020. Eurosurveillance 2020, 25 (10), 2000180. https://doi.org/10.2807/1560-7917.ES.2020.25.10.2000180.

(5) Nishiura, H.; Kobayashi, T.; Miyama, T.; Suzuki, A.; Jung, S.; Hayashi, K.; Kinoshita, R.; Yang, Y.; Yuan, B.; Akhmetzhanov, A. R.; Linton, N. M. Estimation of the Asymptomatic Ratio of Novel Coronavirus Infections (COVID-19). Int J Infect Dis 2020, 94, 154–155. https://doi.org/10.1016/j.ijid.2020.03.020.

(6) Oran, D. P.; Topol, E. J. Prevalence of Asymptomatic SARS-CoV-2 Infection: A Narrative Review. Ann Intern Med 2020, 173 (5), 362–367. https://doi.org/10.7326/M20-3012.

(7) Yu, P.; Zhu, J.; Zhang, Z.; Han, Y. A Familial Cluster of Infection Associated With the 2019 Novel Coronavirus Indicating Possible Person-to-Person Transmission During the Incubation Period. The Journal of Infectious Diseases 2020, 221 (11), 1757–1761. https://doi.org/10.1093/infdis/jiaa077.

(8) Hu, Z.; Song, C.; Xu, C.; Jin, G.; Chen, Y.; Xu, X.; Ma, H.; Chen, W.; Lin, Y.; Zheng, Y.; Wang, J.; Hu, Z.; Yi, Y.; Shen, H. Clinical Characteristics of 24 Asymptomatic Infections with COVID-19 Screened among Close Contacts in Nanjing, China. Sci. China Life Sci. 2020, 63 (5), 706–711. https://doi.org/10.1007/s11427-020-1661-4.

(9) Bai, Y.; Yao, L.; Wei, T.; Tian, F.; Jin, D.-Y.; Chen, L.; Wang, M. Presumed Asymptomatic Carrier Transmission of COVID-19. JAMA 2020, 323 (14), 1406–1407. https://doi.org/10.1001/jama.2020.2565.

(10) Wei, W. E.; Li, Z.; Chiew, C. J.; Yong, S. E.; Toh, M. P.; Lee, V. J. Presymptomatic Transmission of SARS-CoV-2 — Singapore, January 23–March 16, 2020. MMWR Morb Mortal Wkly Rep 2020, 69 (14), 411–415. https://doi.org/10.15585/mmwr.mm6914e1.

(11) Rothe, C.; Schunk, M.; Sothmann, P.; Bretzel, G.; Froeschl, G.; Wallrauch, C.; Zimmer, T.; Thiel, V.; Janke, C.; Guggemos, W.; Seilmaier, M.; Drosten, C.; Vollmar, P.; Zwirglmaier, K.; Zange, S.; Wölfel, R.; Hoelscher, M. Transmission of 2019-NCoV Infection from an Asymptomatic Contact in Germany. New England Journal of Medicine 2020, 382 (10), 970–971. https://doi.org/10.1056/NEJMc2001468.

(12) Linton, N. M.; Kobayashi, T.; Yang, Y.; Hayashi, K.; Akhmetzhanov, A. R.; Jung, S.; Yuan, B.; Kinoshita, R.; Nishiura, H. Incubation Period and Other Epidemiological Characteristics of 2019 Novel Coronavirus Infections with Right Truncation: A Statistical Analysis of Publicly Available Case Data. Journal of Clinical Medicine 2020, 9 (2), 538. https://doi.org/10.3390/jcm9020538.

(13) Backer, J. A.; Klinkenberg, D.; Wallinga, J. Incubation Period of 2019 Novel Coronavirus (2019-NCoV) Infections among Travellers from Wuhan, China, 20-28 January 2020. Euro Surveill 2020, 25 (5). https://doi.org/10.2807/1560-7917.ES.2020.25.5.2000062.

(14) Lauer, S. A.; Grantz, K. H.; Bi, Q.; Jones, F. K.; Zheng, Q.; Meredith, H. R.; Azman, A. S.; Reich, N. G.; Lessler, J. The Incubation Period of Coronavirus Disease 2019 (COVID-19) From Publicly Reported Confirmed Cases: Estimation and Application. Ann Intern Med 2020, 172 (9), 577–582. https://doi.org/10.7326/M20-0504.

(15) Bivins, A.; North, D.; Ahmad, A.; Ahmed, W.; Alm, E.; Been, F.; Bhattacharya, P.; Bijlsma, L.; Boehm, A. B.; Brown, J.; Buttiglieri, G.; Calabro, V.; Carducci, A.; Castiglioni, S.; Cetecioglu Gurol, Z.; Chakraborty, S.; Costa, F.; Curcio, S.; de los Reyes, F. L.; Delgado Vela, J.; Farkas, K.; Fernandez-Casi, X.; Gerba, C.; Gerrity, D.; Girones, R.; Gonzalez, R.; Haramoto, E.; Harris, A.; Holden, P. A.; Islam, Md. T.; Jones, D. L.; Kasprzyk-Hordern, B.; Kitajima, M.; Kotlarz, N.; Kumar, M.; Kuroda, K.; La Rosa, G.; Malpei, F.; Mautus, M.; McLellan, S. L.; Medema, G.; Meschke, J. S.; Mueller, J.; Newton, R. J.; Nilsson, D.; Noble, R. T.; van Nuijs, A.; Peccia, J.; Perkins, T. A.; Pickering, A. J.; Rose, J.; Sanchez, G.; Smith, A.; Stadler, L.; Stauber, C.; Thomas, K.; van der Voorn, T.; Wigginton, K.; Zhu, K.; Bibby, K. Wastewater-Based Epidemiology: Global Collaborative to Maximize Contributions in the Fight Against COVID-19. Environ. Sci. Technol. 2020, 54 (13), 7754–7757. https://doi.org/10.1021/acs.est.0c02388.

(16) Daughton, C. G. Illicit Drugs in Municipal Sewage. In Pharmaceuticals and Care Products in the Environment; ACS Symposium Series; American Chemical Society, 2001; Vol. 791, pp 348–364. https://doi.org/10.1021/bk-2001-0791.ch020.

(17) Choi, P. M.; Tscharke, B. J.; Donner, E.; O’Brien, J. W.; Grant, S. C.; Kaserzon, S. L.; Mackie, R.; O’Malley, E.; Crosbie, N. D.; Thomas, K. V.; Mueller, J. F. Wastewater-Based Epidemiology Biomarkers: Past, Present and Future. TrAC Trends in Analytical Chemistry 2018, 105, 453–469. https://doi.org/10.1016/j.trac.2018.06.004.

(18) Asghar, H.; Diop, O. M.; Weldegebriel, G.; Malik, F.; Shetty, S.; El Bassioni, L.; Akande, A. O.; Al Maamoun, E.; Zaidi, S.; Adeniji, A. J.; Burns, C. C.; Deshpande, J.; Oberste, M. S.; Lowther, S.A. Environmental Surveillance for Polioviruses in the Global Polio Eradication Initiative. The Journal of Infectious Diseases 2014, 210 (suppl_1), S294–S303. https://doi.org/10.1093/infdis/jiu384.

(19) World Health Organization (WHO). Guidelines for Environmental Surveillance of Poliovirus Circulation. 2003.

(20) Hovi, T.; Shulman, L. M.; Avoort, H. V. D.; Deshpande, J.; Roivainen, M.; Gourville, E. M. D. Role of Environmental Poliovirus Surveillance in Global Polio Eradication and Beyond. Epidemiology & Infection 2012, 140 (1), 1–13. https://doi.org/10.1017/S095026881000316X.

(21) Jiehao, C.; Jin, X.; Daojiong, L.; Zhi, Y.; Lei, X.; Zhenghai, Q.; Yuehua, Z.; Hua, Z.; Ran, J.; Pengcheng, L.; Xiangshi, W.; Yanling, G.; Aimei, X.; He, T.; Hailing, C.; Chuning, W.; Jingjing, L.; Jianshe, W.; Mei, Z. A Case Series of Children With 2019 Novel Coronavirus Infection: Clinical and Epidemiological Features. Clinical Infectious Diseases 2020, 71 (6), 1547–1551. https://doi.org/10.1093/cid/ciaa198.

(22) Wang, W.; Xu, Y.; Gao, R.; Lu, R.; Han, K.; Wu, G.; Tan, W. Detection of SARS-CoV-2 in Different Types of Clinical Specimens. JAMA 2020, 323 (18), 1843–1844. https://doi.org/10.1001/jama.2020.3786.

(23) Xiao, F.; Tang, M.; Zheng, X.; Liu, Y.; Li, X.; Shan, H. Evidence for Gastrointestinal Infection of SARS-CoV-2. Gastroenterology 2020, 158 (6), 1831-1833.e3. https://doi.org/10.1053/j.gastro.2020.02.055.

(24) Zhang, J.; Wang, S.; Xue, Y. Fecal Specimen Diagnosis 2019 Novel Coronavirus–Infected Pneumonia. Journal of Medical Virology 2020, 92 (6), 680–682. https://doi.org/10.1002/jmv.25742.

(25) Holshue, M. L.; DeBolt, C.; Lindquist, S.; Lofy, K. H.; Wiesman, J.; Bruce, H.; Spitters, C.; Ericson, K.; Wilkerson, S.; Tural, A.; Diaz, G.; Cohn, A.; Fox, L.; Patel, A.; Gerber, S. I.; Kim, L.; Tong, S.; Lu, X.; Lindstrom, S.; Pallansch, M. A.; Weldon, W. C.; Biggs, H. M.; Uyeki, T. M.; Pillai, S. K. First Case of 2019 Novel Coronavirus in the United States. New England Journal of Medicine 2020. https://doi.org/10.1056/NEJMoa2001191.

(26) Zhang, W.; Du, R.-H.; Li, B.; Zheng, X.-S.; Yang, X.-L.; Hu, B.; Wang, Y.-Y.; Xiao, G.-F.; Yan, B.; Shi, Z.-L.; Zhou, P. Molecular and Serological Investigation of 2019-NCoV Infected Patients: Implication of Multiple Shedding Routes. Emerging Microbes & Infections 2020, 9 (1), 386–389. https://doi.org/10.1080/22221751.2020.1729071.

(27) Wu, Y.; Guo, C.; Tang, L.; Hong, Z.; Zhou, J.; Dong, X.; Yin, H.; Xiao, Q.; Tang, Y.; Qu, X.; Kuang, L.; Fang, X.; Mishra, N.; Lu, J.; Shan, H.; Jiang, G.; Huang, X. Prolonged Presence of SARS-CoV-2 Viral RNA in Faecal Samples. The Lancet Gastroenterology & Hepatology 2020, 5 (5), 434–435. https://doi.org/10.1016/S2468-1253(20)30083-2.

(28) Wölfel, R.; Corman, V. M.; Guggemos, W.; Seilmaier, M.; Zange, S.; Müller, M. A.; Niemeyer, D.; Jones, T. C.; Vollmar, P.; Rothe, C.; Hoelscher, M.; Bleicker, T.; Brünink, S.; Schneider, J.; Ehmann, R.; Zwirglmaier, K.; Drosten, C.; Wendtner, C. Virological Assessment of Hospitalized Patients with COVID-2019. Nature 2020, 581 (7809), 465–469. https://doi.org/10.1038/s41586-020-2196-x.

(29) Tang, A.; Tong, Z.-D.; Wang, H.-L.; Dai, Y.-X.; Li, K.-F.; Liu, J.-N.; Wu, W.-J.; Yuan, C.; Yu, M.-L.; Li, P.; Yan, J.-B. Detection of Novel Coronavirus by RT-PCR in Stool Specimen from Asymptomatic Child, China. Emerg Infect Dis 2020, 26 (6), 1337–1339. https://doi.org/10.3201/eid2606.200301.

(30) Lo, I. L.; Lio, C. F.; Cheong, H. H.; Lei, C. I.; Cheong, T. H.; Zhong, X.; Tian, Y.; Sin, N. N. Evaluation of SARS-CoV-2 RNA Shedding in Clinical Specimens and Clinical Characteristics of 10 Patients with COVID-19 in Macau. Int J Biol Sci 2020, 16 (10), 1698–1707. https://doi.org/10.7150/ijbs.45357.

(31) Han, M. S.; Seong, M.-W.; Kim, N.; Shin, S.; Cho, S. I.; Park, H.; Kim, T. S.; Park, S. S.; Choi, E. H. Viral RNA Load in Mildly Symptomatic and Asymptomatic Children with COVID-19, Seoul, South Korea. Emerg Infect Dis 2020, 26 (10), 2497–2499. https://doi.org/10.3201/eid2610.202449.

(32) Hart, O. E.; Halden, R. U. Computational Analysis of SARS-CoV-2/COVID-19 Surveillance by Wastewater-Based Epidemiology Locally and Globally: Feasibility, Economy, Opportunities and Challenges. Science of The Total Environment 2020, 730, 138875. https://doi.org/10.1016/j.scitotenv.2020.138875.

(33) Walsh, K. A.; Jordan, K.; Clyne, B.; Rohde, D.; Drummond, L.; Byrne, P.; Ahern, S.; Carty, P. G.; O’Brien, K. K.; O’Murchu, E.; O’Neill, M.; Smith, S. M.; Ryan, M.; Harrington, P. SARS-CoV- 2 Detection, Viral Load and Infectivity over the Course of an Infection. Journal of Infection 2020, 81 (3), 357–371. https://doi.org/10.1016/j.jinf.2020.06.067.

(34) Gonzalez, R.; Curtis, K.; Bivins, A.; Bibby, K.; Weir, M. H.; Yetka, K.; Thompson, H.; Keeling, D.; Mitchell, J.; Gonzalez, D. COVID-19 Surveillance in Southeastern Virginia Using Wastewater-Based Epidemiology. Water Research 2020, 186, 116296. https://doi.org/10.1016/j.watres.2020.116296.

(35) Westhaus, S.; Weber, F.-A.; Schiwy, S.; Linnemann, V.; Brinkmann, M.; Widera, M.; Greve, C.; Janke, A.; Hollert, H.; Wintgens, T.; Ciesek, S. Detection of SARS-CoV-2 in Raw and Treated Wastewater in Germany – Suitability for COVID-19 Surveillance and Potential Transmission Risks. Science of The Total Environment 2021, 751, 141750. https://doi.org/10.1016/j.scitotenv.2020.141750.

(36) Ahmed, W.; Angel, N.; Edson, J.; Bibby, K.; Bivins, A.; O’Brien, J. W.; Choi, P. M.; Kitajima, M.; Simpson, S. L.; Li, J.; Tscharke, B.; Verhagen, R.; Smith, W. J. M.; Zaugg, J.; Dierens, L.; Hugenholtz, P.; Thomas, K. V.; Mueller, J. F. First Confirmed Detection of SARS-CoV-2 in Untreated Wastewater in Australia: A Proof of Concept for the Wastewater Surveillance of COVID-19 in the Community. Science of The Total Environment 2020, 728, 138764. https://doi.org/10.1016/j.scitotenv.2020.138764.

(37) La Rosa, G.; Iaconelli, M.; Mancini, P.; Bonanno Ferraro, G.; Veneri, C.; Bonadonna, L.; Lucentini, L.; Suffredini, E. First Detection of SARS-CoV-2 in Untreated Wastewaters in Italy. Science of The Total Environment 2020, 736, 139652. https://doi.org/10.1016/j.scitotenv.2020.139652.

(38) Kumar, M.; Patel, A. K.; Shah, A. V.; Raval, J.; Rajpara, N.; Joshi, M.; Joshi, C. G. First Proof of the Capability of Wastewater Surveillance for COVID-19 in India through Detection of Genetic Material of SARS-CoV-2. Science of The Total Environment 2020, 746, 141326. https://doi.org/10.1016/j.scitotenv.2020.141326.

(39) Medema, G.; Heijnen, L.; Elsinga, G.; Italiaander, R.; Brouwer, A. Presence of SARS-Coronavirus-2 RNA in Sewage and Correlation with Reported COVID-19 Prevalence in the Early Stage of the Epidemic in The Netherlands. Environ. Sci. Technol. Lett. 2020, 7 (7), 511–516. https://doi.org/10.1021/acs.estlett.0c00357.

(40) Lodder, W.; de Roda Husman, de R. SARS-CoV-2 in Wastewater: Potential Health Risk, but Also Data Source. The Lancet Gastroenterology & Hepatology 2020, 5 (6), 533–534. https://doi.org/10.1016/S2468-1253(20)30087-X.

(41) Randazzo, W.; Truchado, P.; Cuevas-Ferrando, E.; Simón, P.; Allende, A.; Sánchez, G. SARS-CoV-2 RNA in Wastewater Anticipated COVID-19 Occurrence in a Low Prevalence Area. Water Research 2020, 181, 115942. https://doi.org/10.1016/j.watres.2020.115942.

(42) Wu, F.; Zhang, J.; Xiao, A.; Gu, X.; Lee, W. L.; Armas, F.; Kauffman, K.; Hanage, W.; Matus, M.; Ghaeli, N.; Endo, N.; Duvallet, C.; Poyet, M.; Moniz, K.; Washburne, A. D.; Erickson, T. B.; Chai, P. R.; Thompson, J.; Alm, E. J. SARS-CoV-2 Titers in Wastewater Are Higher than Expected from Clinically Confirmed Cases. mSystems 2020, 5 (4). https://doi.org/10.1128/mSystems.00614-20.

(43) Nemudryi, A.; Nemudraia, A.; Wiegand, T.; Surya, K.; Buyukyoruk, M.; Cicha, C.; Vanderwood, K. K.; Wilkinson, R.; Wiedenheft, B. Temporal Detection and Phylogenetic Assessment of SARS-CoV-2 in Municipal Wastewater. Cell Reports Medicine 2020, 1 (6), 100098. https://doi.org/10.1016/j.xcrm.2020.100098.

(44) Betancourt, W. Q.; Schmitz, B. W.; Innes, G. K.; Prasek, S. M.; Pogreba Brown, K. M.; Stark, E. R.; Foster, A. R.; Sprissler, R. S.; Harris, D. T.; Sherchan, S. P.; Gerba, C. P.; Pepper, I. L. COVID-19 Containment on a College Campus via Wastewater-Based Epidemiology, Targeted Clinical Testing and an Intervention. Science of The Total Environment 2021, 779, 146408. https://doi.org/10.1016/j.scitotenv.2021.146408.

(45) Gibas, C.; Lambirth, K.; Mittal, N.; Juel, M. A. I.; Barua, V. B.; Brazell, L. R.; Hinton, K.; Lontai, J.; Stark, N.; Young, I.; Quach, C.; Russ, M.; Kauer, J.; Nicolosi, B.; Chen, D.; Akella, S.; Tang, W.; Schlueter, J.; Munir, M. Implementing Building-Level SARS-CoV-2 Wastewater Surveillance on a University Campus. Science of The Total Environment 2021, 146749. https://doi.org/10.1016/j.scitotenv.2021.146749.

(46) Barich, D.; Slonczewski, J. L. Wastewater Virus Detection Complements Clinical COVID-19 Testing to Limit Spread of Infection at Kenyon College. Preprint accessed at medRxiv 2021, 2021.01.09.21249505. https://doi.org/10.1101/2021.01.09.21249505.

(47) Travis, S. A.; Best, A. A.; Bochniak, K. S.; Dunteman, N. D.; Fellinger, J.; Folkert, P. D.; Koberna, T.; Kopek, B. G.; Krueger, B. P.; Pestun, J.; Pikaart, M. J.; Sabo, C.; Schuitema, A. J. Providing a Safe, in-Person, Residential College Experience during the COVID-19 Pandemic. Preprint accessed at medRxiv 2021, 2021.03.02.21252746. https://doi.org/10.1101/2021.03.02.21252746.

(48) Harris-Lovett, S.; Nelson, K.; Beamer, P.; Bischel, H. N.; Bivins, A.; Bruder, A.; Butler, C.; Camenisch, T. D.; Long, S. K. D.; Karthikeyan, S.; Larsen, D. A.; Meierdiercks, K.; Mouser, P.; Pagsuyoin, S.; Prasek, S.; Radniecki, T. S.; Ram, J. L.; Roper, D. K.; Safford, H.; Sherchan, S. P.; Shuster, W.; Stalder, T.; Wheeler, R. T.; Korfmacher, K. S. Wastewater Surveillance for SARS-CoV-2 on College Campuses: Initial Efforts, Lessons Learned and Research Needs. Preprint accessed at medRxiv 2021, 2021.02.01.21250952. https://doi.org/10.1101/2021.02.01.21250952.

(49) Ahmed W; Simpson S; Bertsch P; Bibby K; Bivins A; Blackall L; Bofill-Mas S; Bosch A; Brandao J; Choi P; Ciesielski M; Donner E; D’Souza N; Farnleitner A; Gerrity D; Gonzalez R; Griffith J; Gyawali P; Haas C; Hamilton K; Hapuarachchi C; Harwood V; Haque R; Jackson G; Khan S; Khan W; Kitajima M; Korajkic A; La Rosa G; Layton B; Lipp E; McLellan S; McMinn B; Medema G; Metcalfe S; Meijer W; Mueller J; Murphy H; Naughton C; Noble R; Payyappat S; Petterson S; Pitkanen T; Rajal V; Reyneke B; Roman F; Rose J; Rusinol M; Sadowsky M; Sala-Comorera L; Setoh YX; Sherchan S; Sirikanchana K; Smith W; Steele J; Sabburg R; Symonds E; Thai P; Thomas K; Tynan J; Toze S; Thompson J; Whiteley A; Wong J; Sano D; Wuertz S; Xagoraraki I; Zhang Q; Zimmer-Faust A; Shanks O. Minimizing Errors in RT-PCR Detection and Quantification of SARS-CoV-2 RNA for Wastewater Surveillance. 2021. Preprint accessed at preprints.org https://10.20944/preprints202104.0481.v1

(50) Yang, Q.; Saldi, T; Lasda, E.; Decker, C.; Camille L. Paige; Denise Muhlrad; Patrick K. Gonzales, P.; Fink, M.; Tat, K.; Hager, C.; Davis, J.; Ozeroff, C.; Meyerson, N.; Clark, S.; Fattor, W.; Gilchrist, A;; Barbachano-Guerrero, A.; Worden-Sapper, E.; Wu, S.; Brisson, G.; McQueen, M.; Dowell, R.; Leinwand, L.; Parker, R.; Sawyer, S. Just 2% of SARS-CoV-2-positive individuals carry 90% of the virus circulating in communities. 2021. Preprint accessed at medRxiv doi: https://doi.org/10.1101/2021.03.01.21252250

(51) Bjorkman, K.; Saldi, T.; Lasda, E.; Bauer, L.; Kovarik, J.; Gonzales, P.; Fink, M.; Tat, K.; Hager, C.; Davis, J.; Ozeroff, C.; Brisson, G.; Larremore, D.; Leinwand, L.; McQueen, M.; Parker, R. Higher viral load drives infrequent SARS-CoV-2 transmission between asymptomatic residence hall roommates. 2021. Preprint accessed at medRxiv doi: https://doi.org/10.1101/2021.03.09.21253147

(52) Yang, Q.; Meyerson, N. R.; Clark, S. K.; Paige, C. L.; Fattor, W. T.; Gilchrist, A. R.; Barbachano-Guerrero, A.; Healy, B. G.; Worden-Sapper, E. R.; Wu, S. S.; Muhlrad, D.; Decker, C. J.; Saldi, T. K.; Lasda, E.; Gonzales, P.; Fink, M. R.; Tat, K. L.; Hager, C. R.; Davis, J. C.; Ozeroff, C. D.; Brisson, G. R.; McQueen, M. B.; Leinwand, L. A.; Parker, R.; Sawyer, S. L. Saliva TwoStep for Rapid Detection of Asymptomatic SARS-CoV-2 Carriers. eLife 2021, 10, e65113. https://doi.org/10.7554/eLife.65113.

(53) Cole, D.; Long, S. C.; Sobsey, M. D. Evaluation of F+ RNA and DNA Coliphages as Source-Specific Indicators of Fecal Contamination in Surface Waters. Appl. Environ. Microbiol. 2003, 69 (11), 6507–6514. https://doi.org/10.1128/AEM.69.11.6507-6514.2003.

(54) Boulder County Public Health. Public Health Order 2020-07 Prohibiting Gatherings Involving Persons Aged Between 18 and 22 Years in the City of Boulder and Stay-at-Home for Subject Properties; 2020; Vol. 2020-07.

(55) Crits-Christoph, A.; Kantor, R.: Olm, M.; Whitney, O.; Al-Shayeb, B.; Lou, Y.; Flamholz, A.; Kennedy, L.; Greenwald, H.; Hinkle, A; Hetzel, J.; Spitzer, S.; Koble, J.; Tan, A.; Hyde, F.; Schroth, G.; Kuersten, S; Banfield, J.; Nelson, K. Genome Sequencing of Sewage Detects Regionally Prevalent SARS-CoV-2 Variant. mBio 2021, 12 (1) e02703–20. http://doi.org/10.1128/mBio.02703-20

(56) Shmitz, B.; Innes, G.; Prasek, S.; Betancourt, W.; Stark, E.; Foster, A.; Abraham, G.; Gerba, C.; Pepper, I. Enumerating asymptomatic COVID-19 cases and estimating SARS-CoV-2 fecal shedding rates via wastewater-based epidemiology. 2021. Preprint accessed at medRxiv doi: https://doi.org/10.1101/2021.04.16.21255638

(57) Foladori, P; Cutrupi, F; Segata, N; Manara, S; Pinto, F; Malpei, F; Bruni, L; La Rosa, G. SARS-CoV-2 from faeces to wastewater treatment: What do we know? A review. Science of The Total Environment. 2020. 743, 140444. https://doi.org/10.1016/j.scitotenv.2020.140444.

(58) Minot, S.; Sinha, R.; Chen, J.; Li, H.; Keilbaugh, S.A.; Wu, G.D.; Lewis, J.D.; Bushman, F.D. The human gut virome: inter-individual variation and dynamic response to diet. Genome Research. 2011, 21(10), 1616–1625. https://doi.org/10.1101/gr.122705.111.

(59) Rosario, K.; Symonds, E. M.; Sinigalliano, C.; Stewart, J.; Breitbart, M. Pepper Mild Mottle Virus as an Indicator of Fecal Pollution. Appl. Environ. Microbiol. 2009, 75 (22), 7261–7267. https://doi.org/10.1128/AEM.00410-09.

