## Supplemental Figures for "High-resolution within-sewer SARS-CoV-2 surveillance facilitates informed intervention"

Supplemental Figure 1. Schematic of the sampler design

Supplemental Figure 2. Daily mass of wastewater

Supplemental Figure 3. Daily pH of wastewater

Supplemental Figure 4. Daily total suspended solids (TSS) of wastewater

Supplemental Figure 5. SURV1 Ct values for the extraction blank with a bovine coronavirus recovery spike-in

Supplemental Figure 6. SURV1 Ct values for the positive SARS-CoV-2 control

Supplemental Figure 7. SENB+ Ct values for the A-G standard curves

Supplemental Figure 8. SENB+ concentration (log scale) for the extraction blank with a bovine coronavirus recovery spike-in

Supplemental Figure 9. Performance of the bovine coronavirus spike-in

Supplemental Figure 10. Scatterplot comparing the SARS-CoV-2 nucleocapsid (N) and envelope (E) regions for the (a) SURV1 and (b) SENB+ datasets

Supplemental Figure 11. Comparison of the SARS-CoV-2 concentration with an infection noted within the time window versus those without. The Kruskal-Wallis non-parametric p-value is presented to denote significance (p < 0.05) between the two groups.

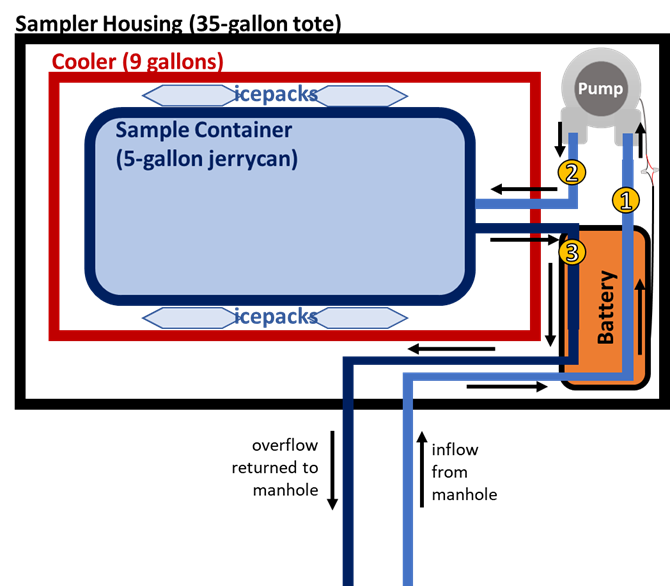

Supplemental Figure 1. Schematic of the sampler design. PVC tubing lines are numbered for easier identification.

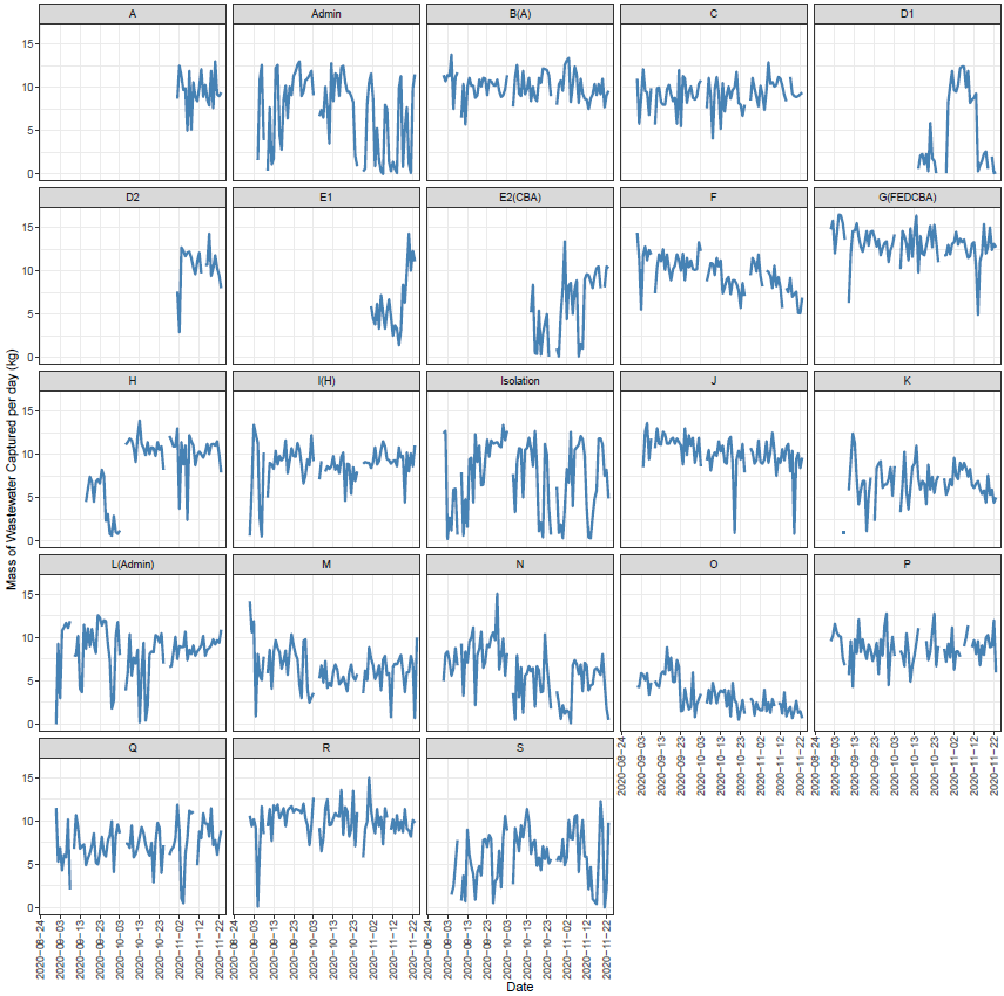

Supplemental Figure 2. Daily mass of wastewater.

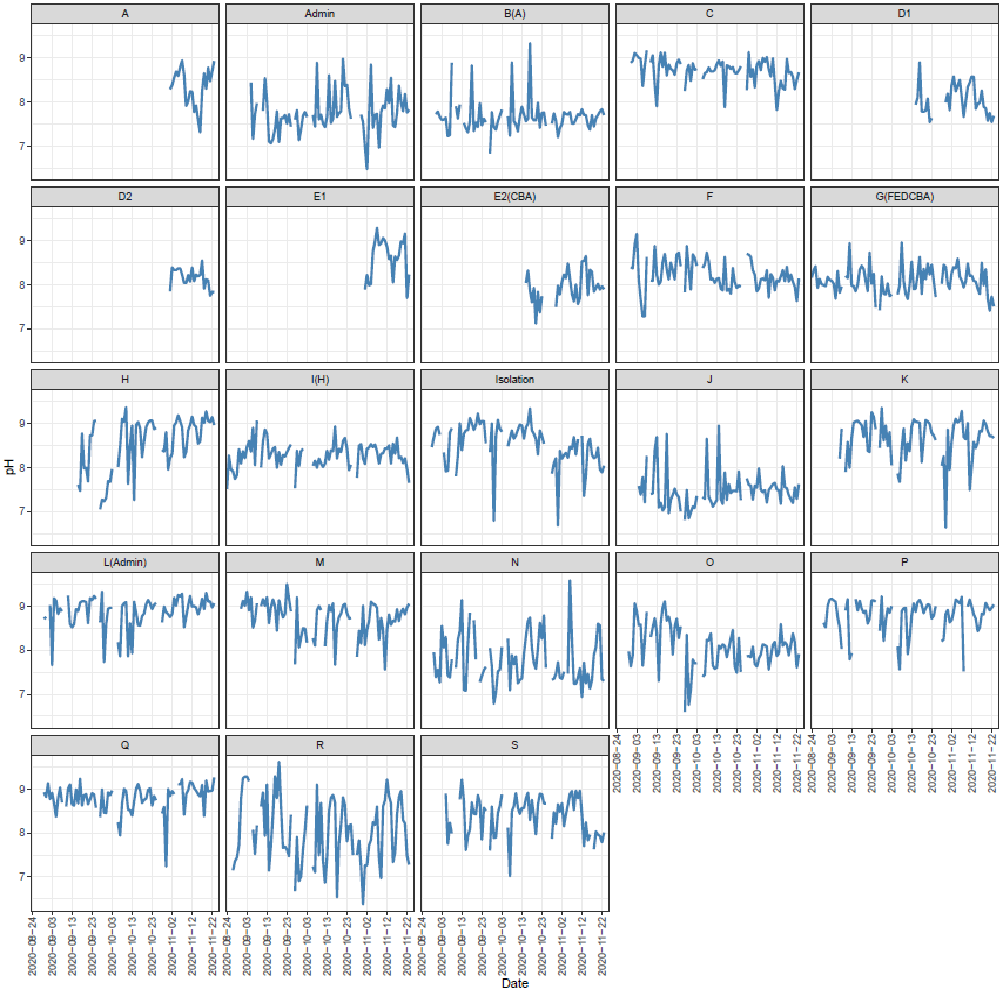
Supplemental Figure 3. Daily pH of wastewater.

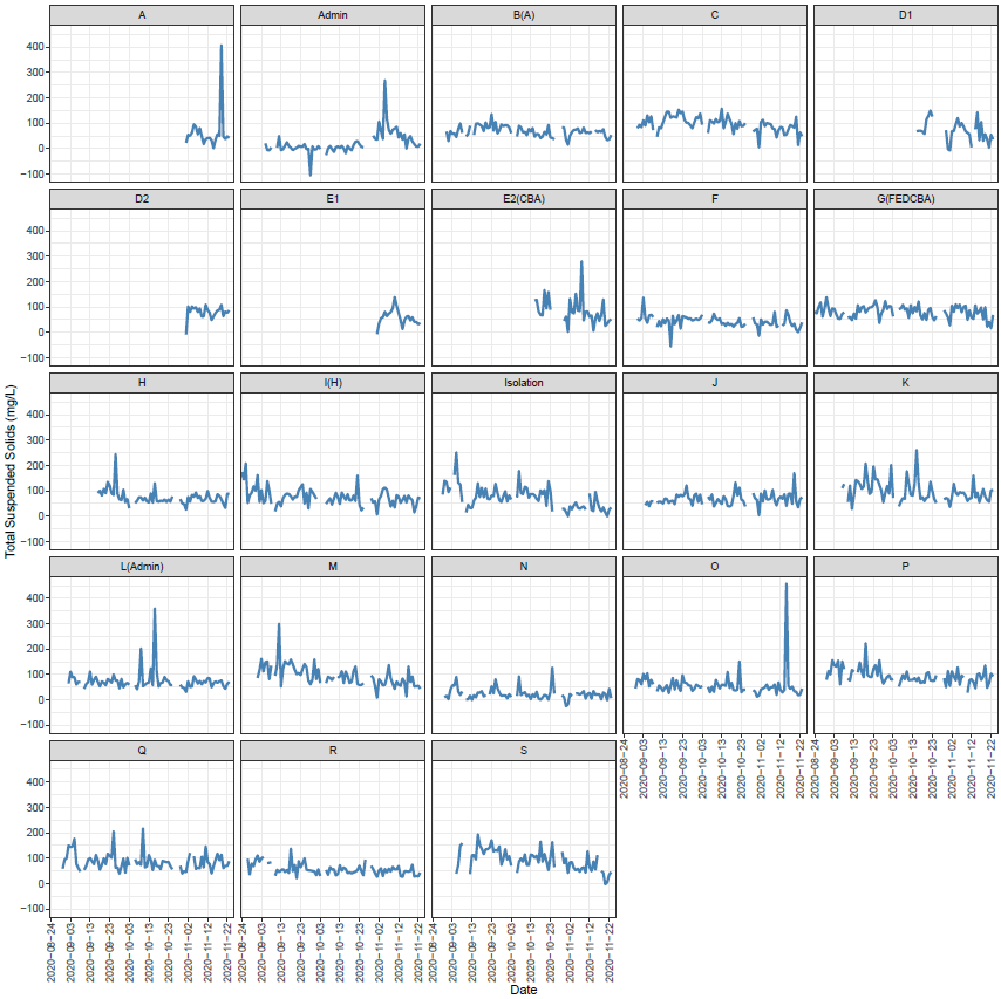

Supplemental Figure 4. Daily total suspended solids (TSS) of wastewater.

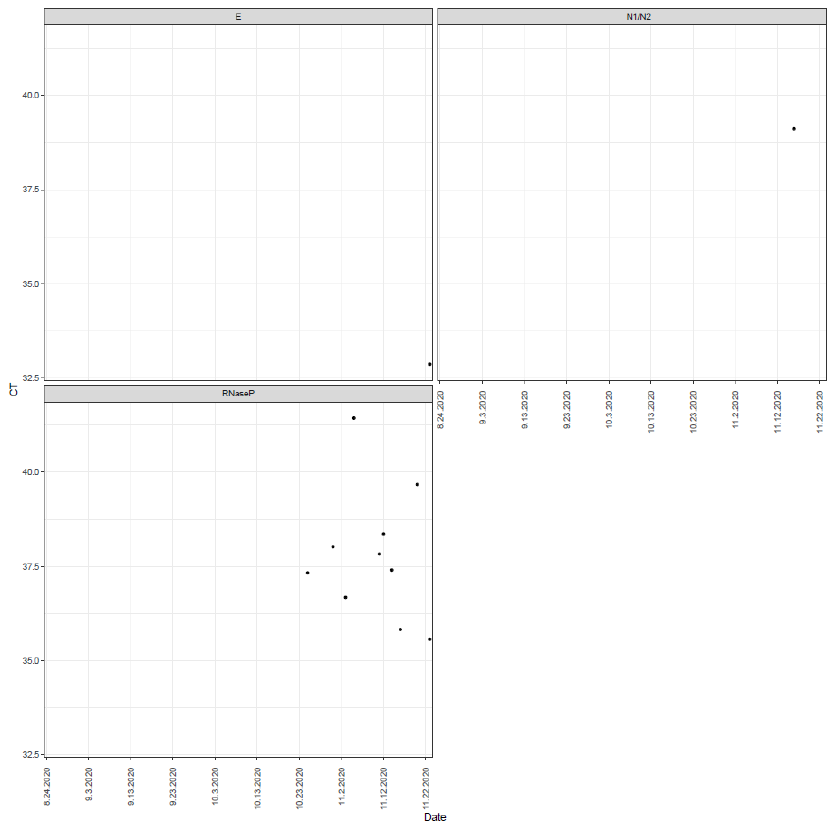

Supplemental Figure 5. SURV1 CT values for the extraction blank with a bovine coronavirus recovery spike-in.

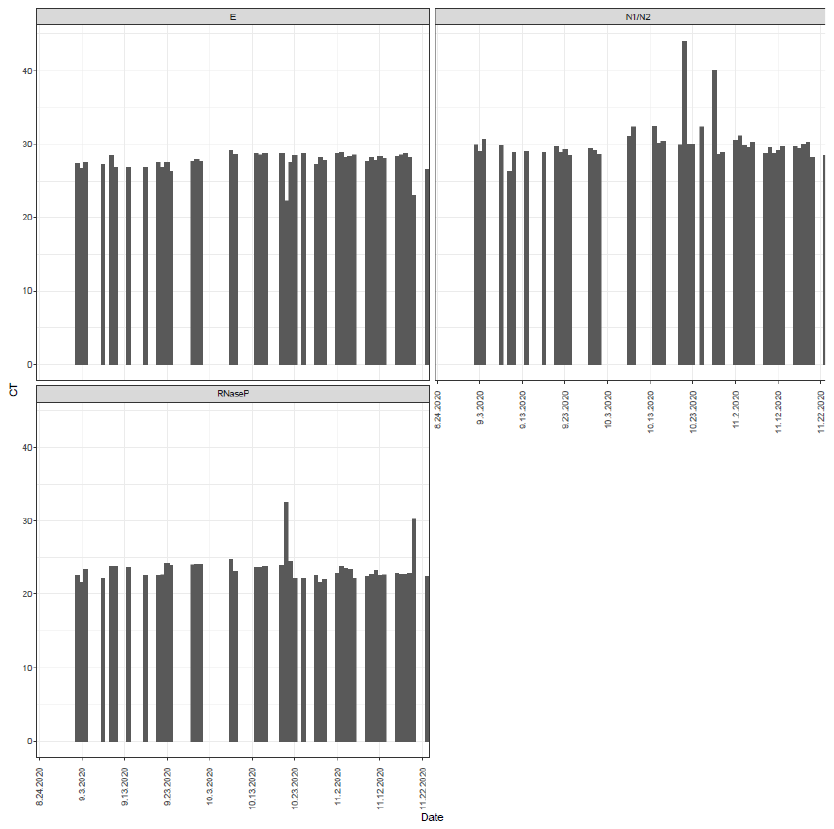
 Supplemental Figure 6. SURV1 CT values for the positive SARS-CoV-2 control.
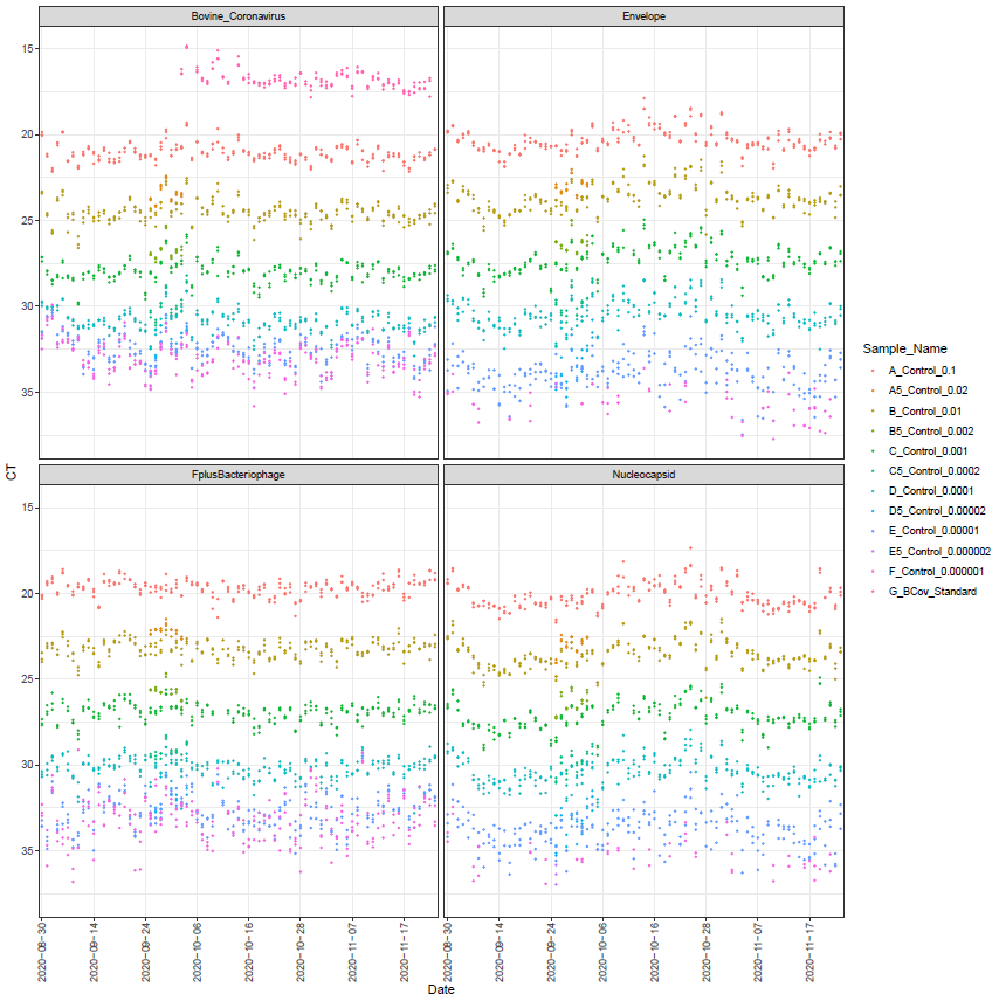

Supplemental Figure 7. SENB+ CT values for the A-G standard curves. A corresponds to 10^5^, 10^6^, and 10^6^ copies per RT-qPCR reaction for SARS-CoV-2, bovine coronavirus, and F+ bacteriophage standard, respectively. The copy number then decreases by one order of magnitude (base 10) until F. F thus corresponds to 1, 10, and 10 copies per RT-qPCR reaction for SARS-CoV-2, bovine coronavirus, and F+ bacteriophage standard, respectively. G corresponds to 1.53×10^7^ copies of bovine coronavirus standard per reaction and was only included on plates when an increased amount of bovine coronavirus spike-in necessitated it.

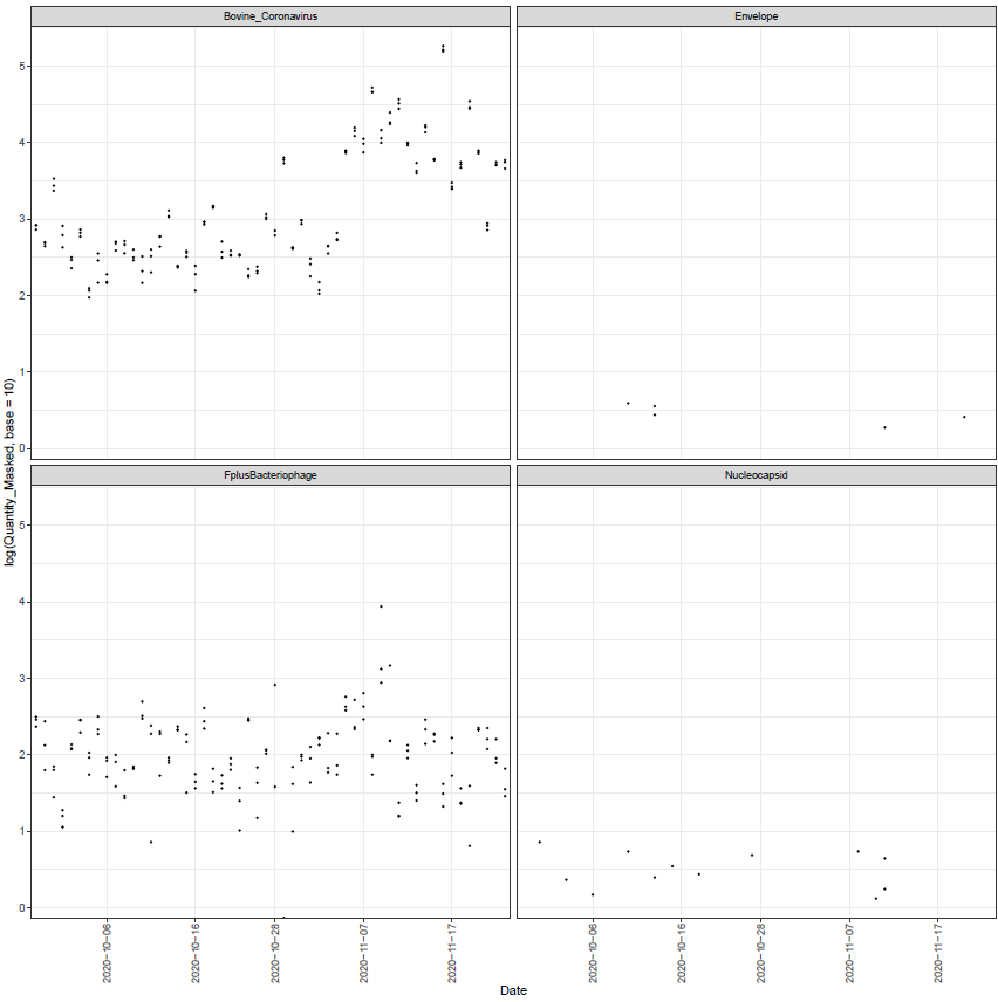

Supplemental Figure 8. SENB+ concentration (log scale) for the extraction blank with a bovine coronavirus recovery spike-in.

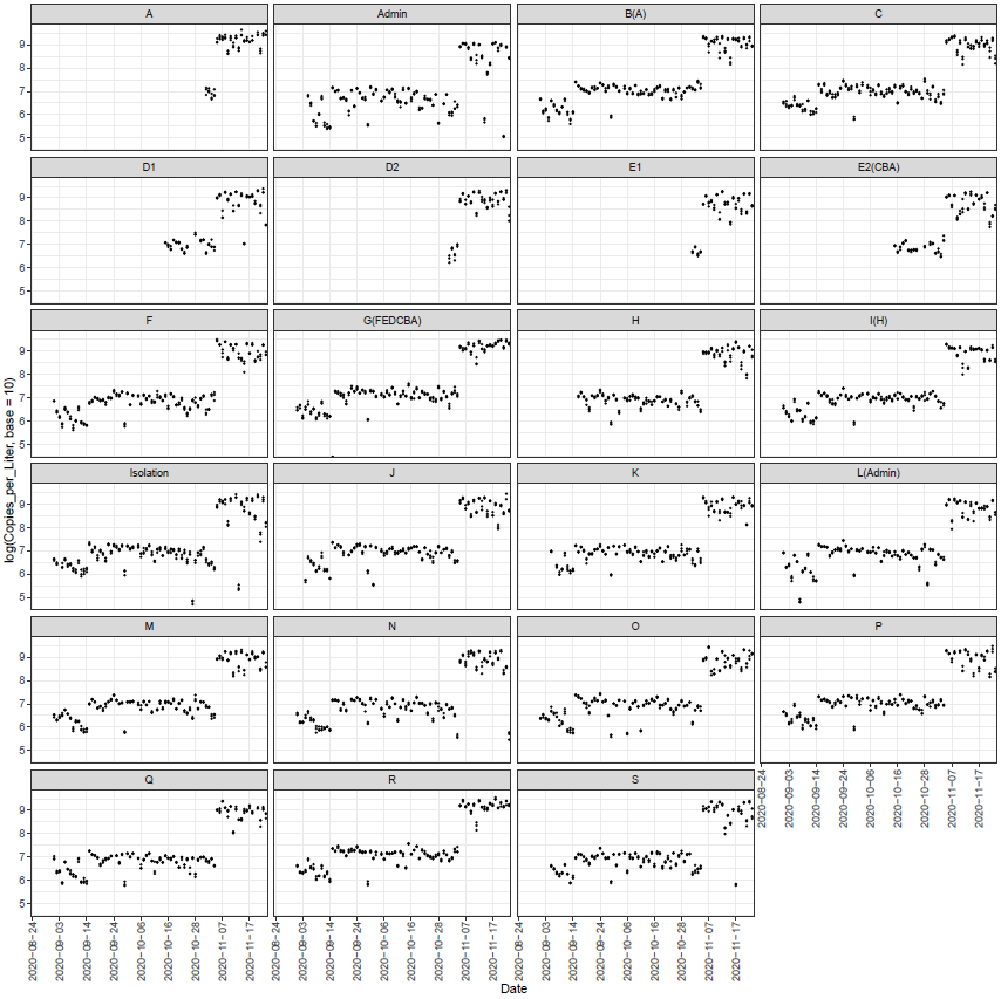

Supplemental Figure 9. Performance of the bovine coronavirus spike-in. Three distinct levels were explored throughout the course of the campaign (2×10^6^, 2×10^7^, and 2×10^9^ copies per liter).

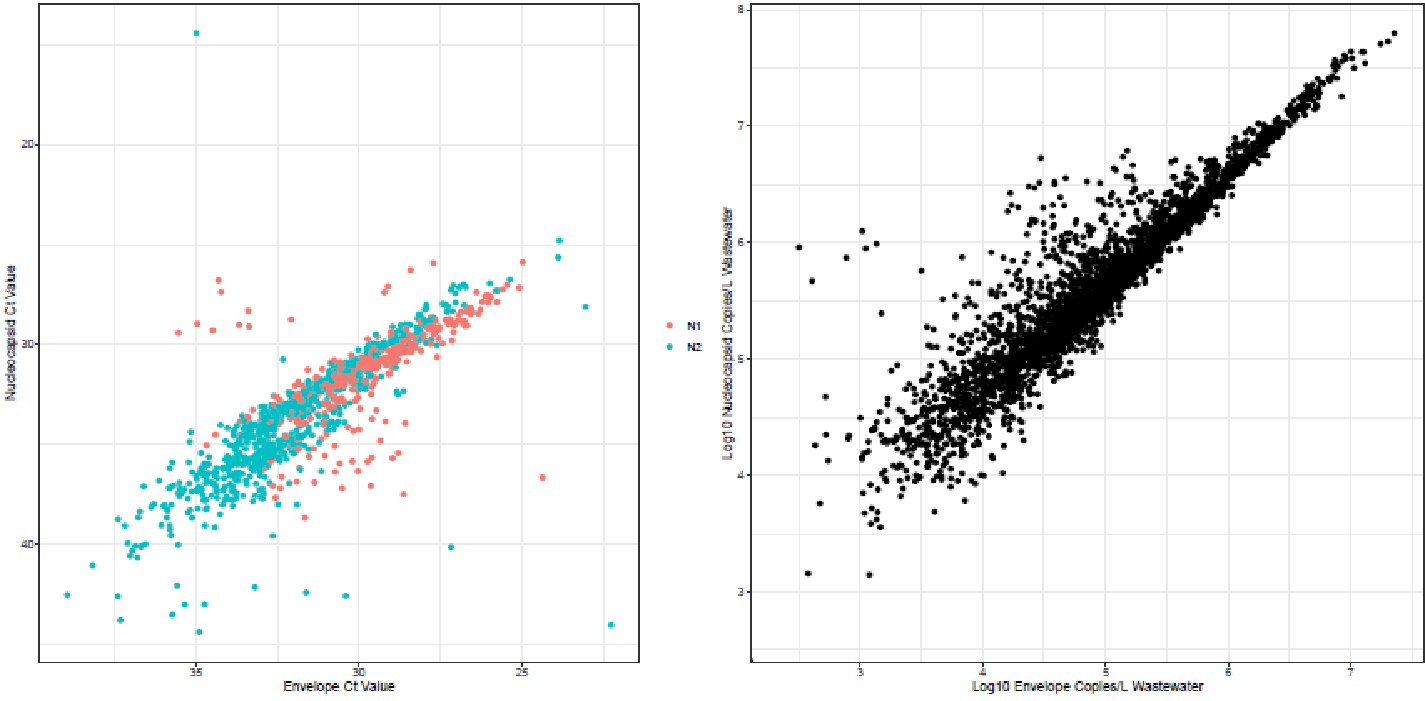

Supplemental Figure 10. Scatterplots comparing the SARS-CoV-2 nucleocapsid (N) and envelope (E) regions for the (left) SURV1 and (right) SENB+ datasets.

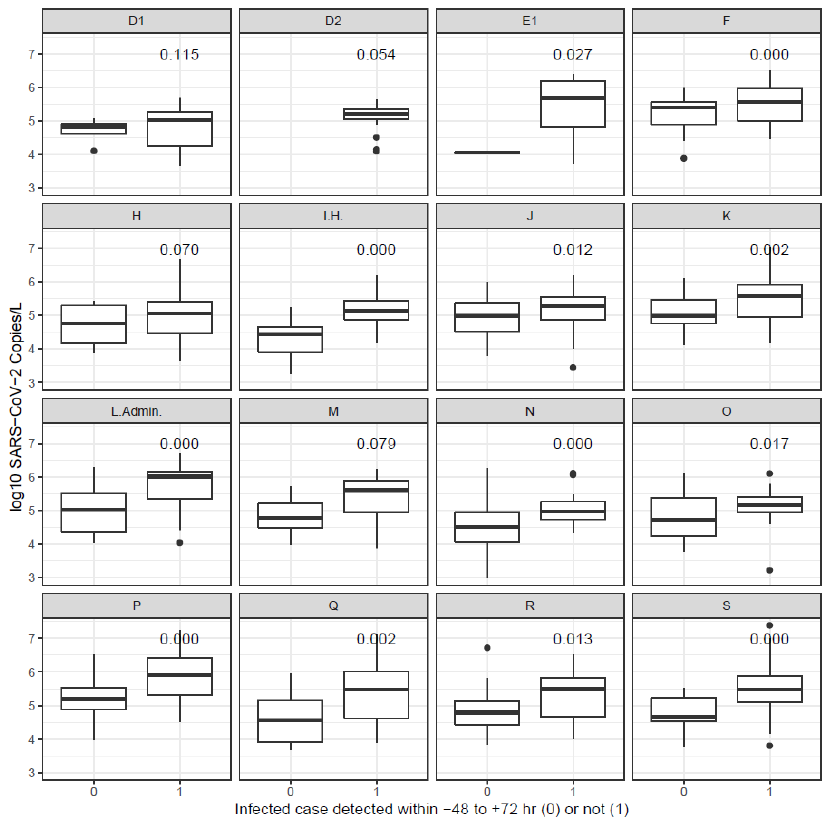

Supplemental Figure 11. Comparison of the SARS-CoV-2 concentration with an infection noted within the time window versus those without. The Kruskal-Wallis non-parametric p-value is presented to denote significance (p < 0.05) between the two groups.
