## Supplemental Script for "High-resolution within-sewer SARS-CoV-2 surveillance facilitates informed intervention": Supplemental_File_05212021.R

```
#======================================================================================================================================
#Install Required Libraries:

#install.packages("ggplot2")
#install.packages("lubridate")
#install.packages("ggplot2")
#install.packages("data.table")
#install.packages("ggrepel")
#install.packages("dplyr")
#install.packages("data.table")
#install.packages("tidyverse")
#install.packages("OpenStreetMap")
#install.packages("corrgram")

#Load Required Libraries:

library(ggplot2); packageVersion("ggplot2")             #At time of implementation, v 3.3.3
library(grid); packageVersion("grid")                   #At time of implementation, v 4.0.3
library(ggfortify); packageVersion("ggfortify")         #At time of implementation, v 0.4.11
library(gridExtra); packageVersion("gridExtra")         #At time of implementation, v 2.3
library(gridGraphics); packageVersion("gridGraphics")   #At time of implementation, v 0.5.1
library(plyr); packageVersion("plyr")                   #At time of implementation, v 1.8.6
library(tidyr); packageVersion("tidyr")                 #At time of implementation, v 1.1.3

#======================================================================================================================================
#######   Read in the qPCR data   ######

qpcr <- read.table("Supplemental File of SENB qPCR Data.txt", header=TRUE)

######    Read in collection Table  ####

collection <- read.table("Supplemental File of Collection Data.txt", header=TRUE)

#Ensure pH is numeric value
collection$pH_Value <- as.numeric(as.character(collection$pH_Value))


###Prepare the Supplemental Mass, pH, and TSS graphics and tables#######

###Supplemental Mass Work#####

#For ease of graphics, create a Mass specific dataframe#
mass_long <- data.frame(collection$Site, as.Date(collection$Date,format = "%m_%d_%y"), collection$Weight_Total_Sample_kg)
colnames(mass_long) <- c('Site', 'Date', 'Mass_kg')

mass_long$Date <- as.character(mass_long$Date)
mass_long$Site <- factor(mass_long$Site)


###Supplemental Figure 3###
#Mass Graph#

pdf("Supplemental_Figure_2.pdf", width = 12, height = 12)

ggplot(mass_long, aes(x = Date, y = Mass_kg, group = Site)) +
  geom_line(color = "steelblue", size = 1) + facet_wrap(~ Site) +
  scale_x_discrete(breaks = levels(factor(mass_long$Date))[c(T, rep(F, 9))]) +
  theme_bw() + ylab("Mass of Wastewater Captured per day (kg)") +
  theme(axis.text.x = element_text(angle = 90, vjust = 0.5, hjust=1))

dev.off()

mass_wide <- spread(mass_long, Site, Mass_kg)

write.table(mass_wide,"Supplemental_Table_3.txt")

pdf("Supplemental_Figure_3.pdf", width = 12, height = 12)

pH_long <- data.frame(collection$Site, as.Date(collection$Date,format = "%m_%d_%y"), collection$pH)
colnames(pH_long) <- c('Site', 'Date', 'pH')
pH_long$Date <- as.character(pH_long$Date)
pH_long$Site <- factor(pH_long$Site)

#pH Graph#

ggplot(pH_long, aes(x = Date, y = pH, group = Site)) +
  geom_line(color = "steelblue", size = 1) + facet_wrap(~ Site) +
  scale_x_discrete(breaks = levels(factor(pH_long$Date))[c(T, rep(F, 9))]) +
  theme_bw() + theme(axis.text.x = element_text(angle = 90, vjust = 0.5, hjust=1))

pH_wide <- spread(pH_long, Site, pH)

dev.off()

write.table(pH_wide,"Supplemental_Table_4.txt")

#TSS

pdf("Supplemental_Figure_4.pdf", width = 12, height = 12)

TSS_long <- data.frame(collection$Site, as.Date(collection$Date,format = "%m_%d_%y"), collection$TSS_Value_gmL)
colnames(TSS_long) <- c('Site', 'Date', 'TSS_g_per_mL')
TSS_long$Date <- as.character(TSS_long$Date)
TSS_long$Site <- factor(TSS_long$Site)

#TSS Graph#

ggplot(TSS_long, aes(x = Date, y = TSS_g_per_mL*1000*1000, group = Site)) +
  geom_line(color = "steelblue", size = 1) + facet_wrap(~ Site) +
  ylab("Total Suspended Solids (mg/L)") +
  scale_x_discrete(breaks = levels(factor(TSS_long$Date))[c(T, rep(F, 9))]) +
  theme_bw() + theme(axis.text.x = element_text(angle = 90, vjust = 0.5, hjust=1))

dev.off()

TSS_wide <- spread(TSS_long, Site, TSS_g_per_mL)

write.table(TSS_wide,"Supplemental_Table_5.txt")


####Supplemental Figure 5######

standard <- qpcr[qpcr$Task == "STANDARD",]
standard$Date <- as.Date(standard$Date,format = "%m/%d/%y")
standard$Date <- factor(standard$Date)
standard$CT <- as.numeric(standard$CT)

pdf("Supplemental_Figure_5.pdf", width = 12, height = 12)

ggplot(standard, aes(x = Date, y = CT, color = Sample_Name, group = Sample_Name)) + 
  geom_point(size=0.5) +
  facet_wrap(~ Target_Name) + scale_y_reverse() +
  scale_x_discrete(breaks = levels(standard$Date)[c(T, rep(F, 9))]) +
  theme_bw() + theme(axis.text.x = element_text(angle = 90, vjust = 0.5, hjust=1))

dev.off()

#####Supplemental Figure 5b#####

pdf("Supplemental_Figure_6.pdf", width = 12, height = 12)

extraction_blank <- qpcr[qpcr$Sample_Name == "Extraction_Blank",]
#Collect only those sites, this is labelled as out "UNKNOWN". I reprocessed the data manually to convert all standards back to standards.

extraction_blank$Date <- as.Date(extraction_blank$Date ,format = "%m/%d/%y")
extraction_blank$Date <- factor(extraction_blank$Date)
extraction_blank$Quantity_Masked <- as.numeric(extraction_blank$Quantity_Masked)


ggplot(extraction_blank, aes(x = Date, y = log(Quantity_Masked,base=10))) +
  geom_point(size=0.5) + facet_wrap(~ Target_Name, ncol=2) + theme_bw() +
  scale_x_discrete(breaks = levels(extraction_blank$Date)[c(T, rep(F, 9))]) + 
  theme(axis.text.x = element_text(angle = 90, vjust = 0.5, hjust=1))

dev.off()

#########################Supplemental Figure 6######################
## Bovine coronavirus recovery ###

bcov.all <- qpcr[qpcr$Target == "Bovine_Coronavirus",]

#Collect only those sites, this is labeled as out "UNKNOWN". 

bcov <- bcov.all[bcov.all$Task == "UNKNOWN",]
bcov <- bcov[bcov$Sample_Name != "G_BCov_Standard",]

bcov$Date <- as.Date(bcov$Date ,format = "%m/%d/%y")
bcov$Date <- factor(bcov$Date)
bcov$Sample_Name <- factor(bcov$Sample_Name)
bcov <- bcov[order(-bcov$Sample_Name),]

pdf("Supplemental_Figure_9.pdf", width = 12, height = 12)

ggplot(bcov, aes(x = Date, y = log(Copies_per_Liter, base = 10))) +
  geom_point(size=0.5) + facet_wrap(~ Sample_Name, ncol=4) + theme_bw() +
  scale_x_discrete(breaks = levels(bcov$Date)[c(T, rep(F, 9))]) + 
  theme(axis.text.x = element_text(angle = 90, vjust = 0.5, hjust=1))

dev.off()

#========================================================================================================================================#
#####Figure 2#####
#========================================================================================================================================#
#Plot the entire dataset for the Fall 2020 semester#

#Subset the data into the target of interest:
envelope <- qpcr[qpcr$Target == "Envelope",]

#Collect only those sites, this is labeled as out "UNKNOWN".
envelope_sites <- envelope[envelope$Task == "UNKNOWN",]

envelope_sparse <- data.frame(envelope_sites$Date, envelope_sites$Sample_Name, envelope_sites$Copies_per_Liter)

mean.df <- aggregate(envelope_sites.Copies_per_Liter ~ envelope_sites.Sample_Name + envelope_sites.Date, envelope_sparse, mean)

#### Incororate the Medical Services Data#########

medical <- as.data.frame(read.table("Supplemental Medical Services Data.txt", header=TRUE))
medical$Date <- as.Date(medical$Date,format = "%m/%d/%y")

##Medical Services by Day and Site###

medical$Date <- factor(medical$Date)
medical.long <- gather(medical, Site, num_positives, A:SUM, factor_key=TRUE)

medical.long.sites <- medical.long[medical.long$Site !="SUM",]


medical.long.sites$Site <- factor(medical.long.sites$Site, levels = rev(c("A","Admin", "B.A.","C","D1","D2","E1","E.CBA.","F","G.FEDCBA.","H","I.H.","Isolation","J","K","L.Admin.","M","N","O","P","Q","R","S", "SUM")))

gg.med_by_site_and_day <- ggplot(medical.long.sites, aes(x = Site, y = Date, fill = num_positives)) +
  geom_tile() + theme_bw() + scale_fill_gradient(low="white", high="black") + 
  scale_y_discrete(limits = levels(medical.long.sites$Date), breaks = levels(medical.long.sites$Date)[c(T, rep(F, 9))]) +
  scale_x_discrete(position = "bottom") + coord_flip() + 
  theme(legend.title = element_blank(), legend.key.size = unit(1, 'cm')) 

##Medical Services by Site####

sum_per_site <- aggregate(medical.long.sites$num_positives, by=list(Group=medical.long.sites$Site), FUN=sum)

population <-read.table("Supplemental File of Population per Structure.txt", header=T)
population.sort <- population[order(population$Site),]

sum_per_site <- sum_per_site[rev(order(sum_per_site$Group)),]

sum_per_site$prevail <- sum_per_site$x/population.sort$Population

sum_per_site$Group <- factor(sum_per_site$Group, levels = rev(c("A","Admin", "B.A.","C","D1","D2","E1","E.CBA.","F","G.FEDCBA.","H","I.H.","Isolation","J","K","L.Admin.","M","N","O","P","Q","R","S", "SUM")))

gg.med_by_site <- ggplot(sum_per_site[sum_per_site$Group != "SUM",], aes(x = Group, y = prevail)) +
  geom_bar(width = 0.7, stat="identity") + coord_flip() +
  theme_bw()

##Medical Services by Day####

medical.long.sum <- medical.long[medical.long$Site =="SUM",]

med.sum.rev <- medical.long.sum[order(medical.long.sum$Date),]

gg.med_by_day <- ggplot(med.sum.rev, aes(x = Date, y = num_positives)) +
  geom_col(width = 0.7) + 
  scale_x_discrete(limits = levels(med.sum.rev$Date), breaks = levels(med.sum.rev$Date)[c(T, rep(F, 9))], position = "bottom") +
  scale_y_reverse(position = "left") + theme_bw()

###Population by Site####

population.sort$Site <- factor(population.sort$Site, levels = rev(c("A","Admin", "B(A)","C","D1","D2","E1","E2(CBA)","F","G(FEDCBA)","H","I(H)","Isolation","J","K","L(Admin)","M","N","O","P","Q","R","S", "SUM")))

gg.pop_by_site <- ggplot(population.sort, aes(x = Site, y = Population)) +
  geom_bar(width = 0.7, stat="identity") + coord_flip() +
  theme_bw()

###Envelope#####

#### Envelope per site and day #####

envelope_sites$Sample_Name <- factor(envelope_sites$Sample_Name, levels = rev(c("A","Admin", "B(A)","C","D1","D2","E1","E2(CBA)","F","G(FEDCBA)","H","I(H)","Isolation","J","K","L(Admin)","M","N","O","P","Q","R","S")))

gg.envelope <- ggplot(envelope_sites, aes(x = as.Date(Date,format = "%m/%d/%y"), y = Sample_Name, fill = log10(Copies_per_Liter))) +
  geom_tile() + theme_bw() + scale_fill_gradient2(midpoint = 5, low="white", mid="blue", high="black") + 
  scale_x_continuous(expand = c(0, 0)) +
  theme(legend.title = element_blank(), legend.key.size = unit(1, 'cm'))

#### Envelope mean per site over the entire campaign #####

envelope_sites$Date <- as.Date(envelope_sites$Date ,format = "%m/%d/%y")
envelope_sites$Sample_Name <- factor(envelope_sites$Sample_Name)
envelope_sites <- envelope_sites[order(-envelope_sites$Sample_Name),]

envelope.na <- envelope_sites
envelope.na$Copies_per_Liter <- replace_na(envelope.na$Copies_per_Liter, 0)
envelope_mean_per_site <- aggregate(envelope.na$Copies_per_Liter, by=list(Group=envelope.na$Sample_Name), FUN=mean)

colnames(envelope_mean_per_site) <- c("Site", "Mean_conc")

envelope_mean_population <- merge(envelope_mean_per_site,population.sort,by="Site")

envelope_mean_population$mean_prevail <- envelope_mean_population$Mean_conc/envelope_mean_population$Population

envelope_mean_population$mean_prevail[2] <- 0

envelope_mean_population$Site <- factor(envelope_mean_population$Site, levels = c("A","Admin", "B(A)","C","D1","D2","E1","E2(CBA)","F","G(FEDCBA)","H","I(H)","Isolation","J","K","L(Admin)","M","N","O","P","Q","R","S"))

gg.mean_per_site <- ggplot(envelope_mean_population, aes(x = Site, y = mean_prevail)) +
  geom_col(width = 0.7) + scale_x_discrete(limits=rev) + ylab("Mean SARS-CoV-2 Envelope Copies/L-person") +
  theme_bw() + coord_flip()

#### Envelope mean per day for all non-isolation sites ######
#### This additionally compares against isolation inclusive #

envelope.na <- envelope_sites
envelope.na$Copies_per_Liter <- replace_na(envelope.na$Copies_per_Liter, 0)
envelope.na.noisolation <- envelope.na[envelope.na$Sample_Name  != "Isolation",]
envelope.na.noisolation.noc <- envelope.na.noisolation[envelope.na.noisolation$Sample_Name  != "C",]

envelope_median_per_date <- aggregate(envelope.na.noisolation.noc$Copies_per_Liter, by=list(Group=envelope.na.noisolation.noc$Date), FUN=median)
envelope_median_per_date2 <- aggregate(envelope.na$Copies_per_Liter, by=list(Group=envelope.na$Date), FUN=median)

colnames(envelope_median_per_date) <- c("Date", "Mean_conc")
colnames(envelope_median_per_date2) <- c("Date", "Mean_conc")

with_iso <- rep("With_Isolation", 87)
without_iso <- rep("Without_Isolation", 87)

w_iso <- cbind(envelope_median_per_date2,with_iso)
colnames(w_iso) <- c("Date", "Mean_conc", "Iso")

wo_iso <- cbind(envelope_median_per_date,without_iso)
colnames(wo_iso) <- c("Date", "Mean_conc", "Iso")

iso_bind <- rbind(w_iso,wo_iso)

iso_bind$Iso <- factor(iso_bind$Iso)

iso_bind$Date <- as.Date(iso_bind$Date ,format = "%m/%d/%y")

gg.median_copies <- ggplot(data = iso_bind, aes(x = Date, y = Mean_conc, fill = Iso)) +
  geom_col(width = 0.7, position = position_dodge()) + ylab("Median Envelope Copies/L-day") + scale_fill_manual(values=c("deepskyblue", "blue")) +
  theme_bw()

#-----------------------------------------------------------------------------------------------------------#
#### Incorporate in the SURV dataset ######

jscbb <- read.table("Supplemental File of SURV qPCR Data.txt", header=TRUE, sep="\t")

jscbb$ID <- factor(jscbb$ID)
jscbb$Target <- factor(jscbb$Target)

jscbb.long <- gather(jscbb, Date, Ct, X8.24.2020:X11.23.2020, factor_key=TRUE)

#Gather the extraction blanks for supplemental display of the run qualities#

jscbb.extraction.blank <- jscbb.long[jscbb.long$ID == "ExtractionBlank",]
jscbb.control <- jscbb.long[jscbb.long$ID == "CONTROL",]

pdf("Supplemental_Figure_7.pdf", width = 12, height = 12)

extraction_blank <- qpcr[qpcr$Sample_Name == "Extraction_Blank",]

jscbb.extraction.blank$Date <- factor(jscbb.extraction.blank$Date)
jscbb.extraction.blank$Ct <- as.numeric(jscbb.extraction.blank$Ct)


ggplot(jscbb.extraction.blank, aes(x = Date, y = Ct)) +
  geom_point(size=1) + facet_wrap(~ Target, ncol=2) + theme_bw() +
  scale_x_discrete(breaks = levels(jscbb.extraction.blank$Date)[c(T, rep(F, 9))]) + 
  theme(axis.text.x = element_text(angle = 90, vjust = 0.5, hjust=1))

dev.off()

#Gather the positive control for supplemental display of the run qualities#

pdf("Supplemental_Figure_8.pdf", width = 12, height = 12)

jscbb.control <- jscbb.long[jscbb.long$ID == "CONTROL",]

jscbb.control$Ct <- as.numeric(jscbb.control$Ct)

ggplot(jscbb.control, aes(x = Date, y = Ct)) +
  geom_bar(stat="identity") + facet_wrap(~ Target, ncol=2) + theme_bw() +
  scale_x_discrete(breaks = levels(jscbb.control$Date)[c(T, rep(F, 9))]) + 
  theme(axis.text.x = element_text(angle = 90, vjust = 0.5, hjust=1))

dev.off()

#Create the ratio of the E target to the RNaseP target#

RNAseP <- jscbb.long[jscbb.long$Target=="RNaseP",]
JSCBB.E <- jscbb.long[jscbb.long$Target=="E",]

ratio <- as.numeric(RNAseP$Ct)-as.numeric(JSCBB.E$Ct)

ratio <- as.numeric(RNAseP$Ct)-as.numeric(JSCBB.E$Ct)
ratio[is.na(ratio)] <- ""
JSCBB.E$ratio <- as.numeric(ratio)

JSCBB.sample <- JSCBB.E[JSCBB.E$Ct != "NOSAMPLE",]
JSCBB.sample.no.blank <- JSCBB.sample[JSCBB.sample$ID != "ExtractionBlank",]
JSCBB.sample.no.extra <- JSCBB.sample.no.blank[JSCBB.sample.no.blank$ID != "CONTROL",]

#Plot the SURV datasett#

gg.jscbb <- ggplot(JSCBB.sample.no.extra, aes(x = Date, y = ID, fill = ratio)) +
  geom_tile() + theme_bw() + scale_y_discrete(limits=rev) +
  scale_x_discrete(limits = levels(JSCBB.sample.no.extra$Date), breaks = levels(JSCBB.sample.no.extra$Date)[c(T, rep(F, 9))], position = "bottom") +
  scale_fill_gradient2(midpoint=5, low="white", mid="red",
                        high="black", space ="Lab" ) + theme(legend.title = element_blank(), legend.key.size = unit(1, 'cm')) 


#Combine into Figure 2; note, some graphics post-processing is conducted in Inkscape to finalize the image #

pdf("Figure_2.pdf", height = 25, width = 20)
grid.arrange(
  grobs = list(gg.median_copies, gg.envelope, gg.mean_per_site, gg.jscbb, 
            gg.pop_by_site, gg.med_by_site_and_day,gg.med_by_site,gg.med_by_day),
  #widths = c(4, 4, 1, 4, 1, 4, 1, 4),
  layout_matrix = rbind(c(1, 1, 1, 1, NA),
                        c(2, 2, 2, 2, 3),
                        c(2, 2, 2, 2, 3),
                        c(4, 4, 4, 4, 5),
                        c(4, 4, 4, 4, 5),
                        c(6, 6, 6, 6, 7),
                        c(6, 6, 6, 6, 7),
                        c(8, 8, 8, 8, NA))
)

dev.off()


#========================================================================================================================================#
#####Figure 3#####
#========================================================================================================================================#
# Plot the isolation data ontop of the number of individuals in the given isolation structure #

isolation <- envelope_sites[envelope_sites$Sample_Name == "Isolation" | envelope_sites$Sample_Name == "C",]
isolation$Date <- as.Date(isolation$Date ,format = "%m/%d/%y")
isolation$Date <- factor(isolation$Date)
isolation$Quantity_Masked <- as.numeric(isolation$Copies_per_Liter)

# Import the number of individuals residing in the isolation structure #

iso_space <- as.data.frame(read.table("Supplemental File of Isolating Population.txt", header=TRUE))
iso_space$Date <- as.Date(iso_space$Date,format = "%m/%d/%y")
iso_space$Date <- factor(iso_space$Date)
iso_space$Hall <- factor(iso_space$Hall)

Scale_factor_for_graphics <- 25

iso_space$RunningTotal <- iso_space$RunningTotal/Scale_factor_for_graphics

pdf("Figure_3.pdf", height = 7/2, width = 9/2)

ggplot() + 
  
  geom_bar(data = iso_space, aes(x = Date, y = RunningTotal, fill = Hall), stat="identity", position=position_dodge()) +
  scale_y_continuous(sec.axis = sec_axis(~.*Scale_factor_for_graphics, name = "Residents in Isolation")) + scale_fill_manual(values=c("deepskyblue", "brown1")) +
  
  geom_point(data = isolation, aes(x = Date, y = log(Copies_per_Liter, base = 10), color = Sample_Name), size = 1) +
  theme_bw() + scale_color_manual(values=c("red", "blue")) +
  scale_x_discrete(breaks = levels(iso_space$Date)[c(T, rep(F, 19))]) + 
  theme(axis.text.y.left = element_text(color="purple"),
        axis.title.y.left = element_text(colour = "purple"),
        axis.line.y.left = element_line(colour = "purple")
  ) +
  ylab("Log10 Copies of SARS-CoV-2 Envelope/L") + theme(legend.position = "none")
dev.off()


#######################Supplmental Nucleo Figure#################################

nucleo <- qpcr[qpcr$Target == "Nucleocapsid",]
nucleo_sites <- nucleo[nucleo$Task == "UNKNOWN",]
nucleo_t <- nucleo_sites

###Nucelo#####

nucleo_t$Sample_Name <- factor(nucleo_t$Sample_Name, levels = rev(c("A","Admin", "B(A)","C","D1","D2","E1","E2(CBA)","F","G(FEDCBA)","H","I(H)","Isolation","J","K","L(Admin)","M","N","O","P","Q","R","S")))
gg.nucleo <- ggplot(nucleo_t, aes(x = as.Date(Date,format = "%m/%d/%y"), y = Sample_Name, fill = log10(Copies_per_Liter))) +
  geom_tile() + theme_bw() + scale_fill_gradient2(midpoint = 5, low="white", mid="blue", high="black") + 
  scale_x_continuous(expand = c(0, 0)) +
  theme(legend.title = element_blank(), legend.key.size = unit(1, 'cm'))

nucleo_t$Date <- as.Date(nucleo_t$Date ,format = "%m/%d/%y")
nucleo_t$Sample_Name <- factor(nucleo_t$Sample_Name)
nucleo_t <- nucleo_t[order(-nucleo_t$Sample_Name),]

jscbb.long.noempty <- jscbb.long[jscbb.long$Target != "",]
jscbb.long.noempty.ID <-jscbb.long.noempty[jscbb.long.noempty$Target != "ExtractionBlank",]
jscbb.long.sample <- jscbb.long.noempty.ID[jscbb.long.noempty.ID$Ct != "NOSAMPLE",]
jscbb.data_wide <- spread(jscbb.long.sample, Target, Ct)

jscbb.data_wide.G <- jscbb.data_wide[jscbb.data_wide$ID=="G(FEDCBA)",]
jscbb.data_wide.G$Date

new_id_N1N2 <- mapvalues(jscbb.data_wide$Date, 
                    from = c( "X8.26.2020","X8.27.2020","X8.28.2020","X8.29.2020","X8.30.2020","X8.31.2020","X9.1.2020","X9.2.2020","X9.3.2020","X9.4.2020","X9.5.2020",
                              "X9.6.2020","X9.7.2020","X9.8.2020","X9.10.2020","X9.11.2020","X9.12.2020","X9.13.2020","X9.14.2020","X9.15.2020","X9.16.2020","X9.17.2020",
                              "X9.18.2020","X9.19.2020","X9.20.2020","X9.21.2020","X9.22.2020","X9.23.2020","X9.24.2020","X9.25.2020","X9.26.2020","X9.27.2020","X9.28.2020", 
                              "X9.29.2020","X9.30.2020","X10.1.2020","X10.2.2020","X10.3.2020","X10.6.2020","X10.7.2020","X10.8.2020","X10.9.2020","X10.10.2020","X10.11.2020",
                              "X10.12.2020","X10.13.2020","X10.14.2020","X10.15.2020","X10.16.2020","X10.17.2020","X10.18.2020","X10.19.2020","X10.20.2020","X10.21.2020","X10.22.2020",
                              "X10.23.2020","X10.24.2020","X10.25.2020","X10.28.2020","X10.29.2020","X10.30.2020","X10.31.2020","X11.1.2020","X11.2.2020","X11.3.2020","X11.4.2020",
                              "X11.5.2020","X11.6.2020","X11.7.2020","X11.8.2020","X11.9.2020","X11.10.2020","X11.11.2020","X11.12.2020","X11.13.2020","X11.14.2020","X11.15.2020",
                              "X11.16.2020","X11.17.2020","X11.18.2020","X11.19.2020","X11.20.2020","X11.21.2020","X11.22.2020","X11.23.2020"),
                    to = c(rep("N1",33),rep("N2",52)))

jscbb.data_wide$Date <- new_id_N1N2

gg.SURV.NE <- ggplot(jscbb.data_wide, aes(x = as.numeric(E) ,
                     y =  as.numeric(N1), color = Date)) + geom_point() + theme_bw() + scale_y_reverse() + scale_x_reverse() +
                     ylab("Nucleocapsid Ct Value") + xlab("Envelope Ct Value") + theme(legend.title = element_blank())


compareEN <- qpcr[qpcr$Task == "UNKNOWN",]

compareEN.Envelope <- compareEN[compareEN$Target == "Envelope",]
compareEN.Nucleo <- compareEN[compareEN$Target == "Nucleocapsid",]

EN_copies <- cbind(compareEN.Envelope$Copies_per_Liter,compareEN.Nucleo$Copies_per_Liter)
colnames(EN_copies) <- c("Envelope","Nucleo")

gg.SENB.NE <- ggplot(EN_copies, aes(x = log(as.numeric(Envelope), base=10) ,
                            y =  log(as.numeric(Nucleo),base=10))) + geom_point() + theme_bw() + 
  ylab("Log10 Nucleocapsid Copies/L Wastewater") + xlab("Log10 Envelope Copies/L Wastewater") + theme(legend.title = element_blank())


pdf("Supplemental_Figure_10.pdf", height = 8, width = 16)

grid.arrange(
  grobs = list(gg.SURV.NE, gg.SENB.NE),
  layout_matrix = rbind(c(1,2))
)

dev.off()

#========================================================================================================================================#
#####Figure 4#####
#========================================================================================================================================#

#Plot the F+ Bacteriophage data#

fplus <- qpcr[qpcr$Target == "FplusBacteriophage",]
fplus_sites <- fplus[fplus$Task == "UNKNOWN",]

fplus_sites$Date <- as.Date(fplus_sites$Date ,format = "%m/%d/%y")
fplus_sites$Date <- factor(fplus_sites$Date)

fplus_t <- fplus_sites

fplus_t$Sample_Name <- factor(fplus_t$Sample_Name, levels = rev(c("A","Admin", "B(A)","C","D1","D2","E1","E2(CBA)","F","G(FEDCBA)","H","I(H)","Isolation","J","K","L(Admin)","M","N","O","P","Q","R","S")))

pdf("Figure_4.pdf", height = 10, width = 10)

ggplot(fplus_t, aes(x = Date, y = Sample_Name, fill = log10(Copies_per_Liter))) +
  geom_tile() + theme_bw() + ylab("Sites") +
  scale_x_discrete(limits = levels(fplus_t$Date), breaks = levels(fplus_t$Date)[c(T, rep(F, 19))], position = "bottom") +
  scale_fill_gradient2(midpoint=6.5, low="white", mid="dark orange",
                       high="black", space ="Lab" ) + theme(legend.title = element_blank(), legend.key.size = unit(0.5, 'cm')) 

dev.off()
```
